# Deleterious drugs in COVID-19: a rapid systematic review and meta-analysis

**DOI:** 10.1101/2021.09.17.21262724

**Authors:** Michael Holder, Catherine Heeney, Stephen Malden, Uditha Perera, Aziz Sheikh

**Affiliations:** Usher Institute, University of Edinburgh, Edinburgh, United Kingdom

## Abstract

**Background:** Concerns have been expressed about a number of drugs that potentially worsen outcomes in patients with COVID-19. We sought to identify all potentially deleterious drug groups in COVID-19 and critically assess the underpinning strength of evidence pertaining to the harmful effects of these drugs.

**Methods and findings:** We performed a rapid systematic review, searching Medline, Embase and two COVID-19 portfolios (WHO COVID-19 database and NIH iSearch COVID-19 portfolio) for papers and preprints related to primary studies investigating drugs identified as potentially deleterious. Primary outcomes were direct measures of susceptibility to infection, disease severity and mortality. Study quality was assessed using the National Heart, Lung, and Blood Institute quality assessment tools. Random-effects meta-analyses were used for data synthesis with further subgroup analyses where possible for specific outcome, study design, statistical adjustment and drug groups when two were combined. Sensitivity analyses were performed by removing any studies at high risk of bias and by publication status.

49 observational studies (15 peer-reviewed papers and 34 preprints) reported primary outcomes for eight drug groups hypothesised to be deleterious. Meta-analysis showed that acute inpatient corticosteroid use was associated with increased mortality (OR 2.22, 95% CI 1.26-3.90), however this result appeared to have been biased by confounding via indication. One subgroup analysis indicated an association between immunosuppressant use and susceptibility to COVID-19 among case control and cross-sectional studies (OR 1.29, 95% CI 1.19-1.40) but this was not found with cohort studies (OR 1.11, 95% CI 0.86-1.43). Studies which adjusted for multiple confounders showed that people taking angiotensin-converting-enzyme inhibitors (ACEIs) or angiotensin-II-receptor blockers (ARBs) required a lower level of care (OR 0.85, 95% CI 0.74-0.98). Furthermore, studies which combined these two drug groups in their analysis demonstrated an association with a lower mortality (OR 0.68, 95% CI 0.55-0.85).

**Conclusions:** We found minimal high quality or consistent evidence that any drug groups increase susceptibility, severity or mortality in COVID-19. Converse to initial hypotheses, we found some evidence that regular use of ACEIs and ARBs prior to infection may be effective in reducing the level of care required, such as requiring intensive care, in patients with COVID-19.

## Introduction

The ongoing Coronavirus 2019 (COVID-19) pandemic has already had a profound global impact. As of 3^rd^ March 2021, there had been over 114 million confirmed cases and over 2.5 million deaths worldwide (1). SARS-CoV-2 is one of seven coronaviruses known to infect humans (2). It is, together with severe acute respiratory syndrome coronavirus (SARS-CoV) and the Middle East respiratory syndrome coronavirus (MERS-CoV), also one of three zoonotic human betacoronaviruses to have emerged in the last 20 years (3). The genome sequence of SARS-CoV-2 was found to be 79% similar to SARS-CoV (4); both viruses enter human cells via angiotensin-converting enzyme 2 (ACE2), a factor linked to human-to-human transmission (5). ACE2 received considerable attention following its discovery as the entry for SARS-CoV, and has previously been purported to play a protective role in disease, as opposed to the deleterious role of angiotensin-converting enzyme (ACE) (6). Conversely, some hypothesized that drugs which upregulate ACE2 may increase the risk of severe SARS-CoV-2 infection (7).

Approximately five percent of known COVID-19 cases require admission to an intensive care unit (ICU) (8). Many of these patients develop acute respiratory distress syndrome (ARDS) and require mechanical ventilation (MV) or other respiratory support (9). Cytokine storms (a large release of pro-inflammatory proteins) have been implicated in the development of ARDS, as well as multi-organ failure, with a correlation between cytokine levels and mortality (10).

Since the beginning of the pandemic, both expert opinion and small-scale observational studies have contributed to a number of drug groups being postulated to alter the progression of the disease, whether positively or negatively. At various times, conflicting advice regarding non-steroidal anti-inflammatory drugs (NSAIDs) (11–13), corticosteroids (14, 15) and other immunosuppressants (16–18), ACE Inhibitors (ACEIs) and Angiotensin Receptor Blockers (ARBs) (19) have been reported in the scientific literature and media, leading to considerable confusion for both patients and healthcare professionals.

Especially during the early stages of the pandemic, most of the advice stemmed from outcomes in other respiratory infections including severe acute respiratory syndrome (SARS) (20, 21) and Middle East respiratory syndrome (MERS) (14) or hypotheses based on the mechanism of viral entry into cells (19, 22, 23) and general effects on immunity (17, 18). In addition, many scientific papers have been published prior to peer review to speed up information sharing, which has led to questions surrounding the scientific validity of some of these findings (24).

We aimed to identify, critically appraise and synthesise the evidence on drugs which may be deleterious in COVID-19.

## Methods

This study is reported as per PRISMA guidelines on the reporting of systematic reviews (25). In response to the exponential rate at which new research on this topic is being published, a rapid review methodology was adopted following the Cochrane rapid review protocol (26) with the aim of producing reliable results in a timely fashion.

### Search strategy

As the availability of studies increased, our search strategy evolved iteratively to capture as much as possible of the available evidence base. This study consisted of a three-stage search strategy involving literature from four databases using the PECOS format (Population, Exposure, Comparison, Outcome, Study design): Embase, Medline, World Health Organization (WHO) COVID-19 Database (27) and National Institutes for Health (NIH) iSearch COVID-19 Portfolio (28). The searches were performed in April 2020 and included all papers between December 2019 and April 2020. We first searched the Medline and Embase databases simultaneously through Ovid, using a combination of Medical Subject Headings (MESH) and keywords, to capture papers related to the drugs already identified as potentially harmful in exploratory searches, as well as a string to identify new drugs. (Appendix 1: search terms). The primary goal of this search was to amass a list of drugs hypothesized as potentially harmful in COVID-19 in order to inform the subsequent searches.

The second search was performed on the WHO COVID-19 Database (27), a database aimed at compiling all the available publications on COVID-19. This search used terms to capture evidence related to all drugs identified prior to the second search.

Due to the rapidity of emerging evidence, we felt we could not rely solely on peer-reviewed publications as this would severely limit our findings and we decided to include preprints a priori, which at the time were the primary source of information available on COVID-19 epidemiology and pharmacovigilance (29). The third and final search was, therefore, undertaken for preprints on the NIH iSearch COVID-19 Portfolio database, which included Research Square, medRxiv, chemRxiv, arXiv, bioRxiv and Social Science Research Network. This search was screened for primary data only.

### Screening

As per the Cochrane rapid review guidelines (26), all abstracts were screened by the first reviewer (MH). A second reviewer screened any excluded papers (SM and CH), while a third reviewer helped to settle any disagreements (UP). All studies included were then screened as a full text in the same manner, with all included full texts being moved on to extraction. Finally, this process was repeated for reference lists from all relevant systematic reviews identified from the searches.

The first two searches were initially screened with the aim of capturing a comprehensive list of drugs hypothesized to be harmful and therefore included opinion pieces. These searches were then screened a second time for papers with primary data using a second set of inclusion and exclusion criteria (Appendix 2: inclusion & exclusion criteria). The third search was screened only for primary data using the second set of criteria.

### Data extraction

Data extraction was performed using a tailored data extraction template designed to capture relevant data pertaining to study design, setting, demographics and findings (Appendix 5). Data from each study were extracted by the first reviewer (MH) and checked independently by a second reviewer (UP).

### Quality assessment

Quality assessment was performed using the National Heart, Lung, and Blood Institute (NHLBI) checklists (30). We chose this tool for three reasons: it is a validated and reliable tool; it contains checklists to assess a large range of observational study designs;(13) and it is relatively simple and quick to administer in comparison to other quality assessment tools, which aligned with the rationale for adopting a rapid review methodology. Two of the four reviewers (MH and SM/CH/UP) checked the quality of each study against the relevant checklist and reached consensus through discussion when in disagreement.

No formal assessment of strength of evidence by outcome was undertaken; however, we considered our findings in line with the key aspects of the grading of recommendations assessment, development and evaluation (GRADE) (31).

### Outcome definitions

Drug group analysis was split into three categories: susceptibility to infection; severity of disease; and mortality. Susceptibility was defined as testing positive, clinical diagnosis or hospitalisation (if compared with untested individuals) as these all compared case numbers against those without the disease. Severity of disease was split further where possible into either a diagnostic index or level of care. Diagnostic index of severity was defined as any combination of factors used to differentiate severe or critical disease from mild or moderate, or diagnostic criteria for ARDS. Level of care was defined as hospitalisation (if compared with community care of COVID-19 cases), ventilation (mechanical or non-invasive ventilation), ICU care or mortality in combination with other outcomes including remaining as an inpatient. Mortality was defined as any measure of mortality outside of or within hospital, either within a set time frame or at any point during data collection.

Although hospitalisation rates would normally imply more severe disease than simply testing positive, in many countries testing was largely only available in hospitals (32–34). If a study compared those admitted to hospital due to COVID-19 with untested individuals, this was considered analogous with testing positive for the purpose of analysis. However, if the control group contained positive cases tested in the community, then this was considered as requiring a higher level of care.

Due to the common pathway affected by both ACEIs and ARBs, and the recommendation against co-prescription (35), these drug groups were combined in analyses. Henceforth, when combined, ACEIs and ARBs will be referred to as renin-angiotensin-aldosterone blockers (RAASBs).

For the purposes of this review, we considered immunosuppressants as a class of drugs used primarily to suppress the human immune system. Most included studies investigated immunosuppressants in this manner, although some studies analysed individual immunosuppressants. Where corticosteroids were grouped together with other immunosuppressants, these studies were included here.

### Narrative synthesis

For groups of outcomes for a particular drug group that had fewer than four effect estimates with calculable and relevant odds ratios (ORs), these were synthesised narratively. Study description tables were used to summarise study characteristics and compare findings.

### Statistical analysis

Any drug groups with a sufficient number of comparable studies were synthesised using meta-analyses. Considering the implications of random-effects models on statistical power (36), outcomes were deemed eligible for meta-analysis if more than four studies were identified that investigated the same drug group. When sufficient numbers of results were available, these were then run with sub-group analyses to attempt to address any heterogeneity in the results and investigate which confounding factors may have affected findings. The subgroup analyses included study design, level of adjustment for confounders, acute doses or regular users, the two subsets of disease severity defined above and whether ACEIs and ARBs were analysed together or separately. Additionally, preprint studies that had not yet been formally peer-reviewed were also removed during sensitivity analyses.

We included studies that either reported an OR, relative risk (RR) or the raw data that allowed the calculation of OR in the meta-analyses. If available, we used an adjusted OR, otherwise we used an unadjusted OR, either reported in the paper or manually calculated. If the calculated OR exhibited a different result from the conclusion reached by the paper, this was not included in the results.

If more than one outcome was reported, these were all included if they were in different categories (i.e. severe disease and mortality). If multiple outcome subsets were reported within the same categories, the outcome with the highest number of patients in the exposed group, and therefore greatest weight, was used. The only deviation from this was within the severity of disease category, where the diagnostic index would take precedence to reduce any potential selection bias for increased level of care.

We used MetaXL (Version 5.3; EpiGear International Pty Ltd) to run random-effects meta-analyses and create forest plots and funnel plots. Results were reported with a 95% confidence interval (CI). We used Cochrane’s Q and I^2^ test to assess for heterogeneity. An I^2^ <30% was considered low heterogeneity, ≥30% but <50% was considered moderate and ≥50% was considered high heterogeneity (37). Sensitivity analyses were performed on all models by removing each study to assess their risk of bias on the pooled effect estimates and heterogeneity. Publication bias was assessed for any meta-analyses that contained 10 or more studies by visually inspecting funnel plots for asymmetry.

## Results

### Eligible studies identified

From the first two searches, we identified eight drug groups that were hypothesized to be deleterious in COVID-19 from 178 peer-reviewed publications comprising ACEIs, ARBs, corticosteroids, Immunosuppressants, mineralocorticoid receptor antagonists (MCRAs), NSAIDs, Statins and Thiazolidinediones (TZDs).

From the combined total of papers from all three searches, we identified 63 papers with primary data related to these drugs, with 49 measuring at least one primary outcome included in the study (Fig 1).

**Fig 1.**
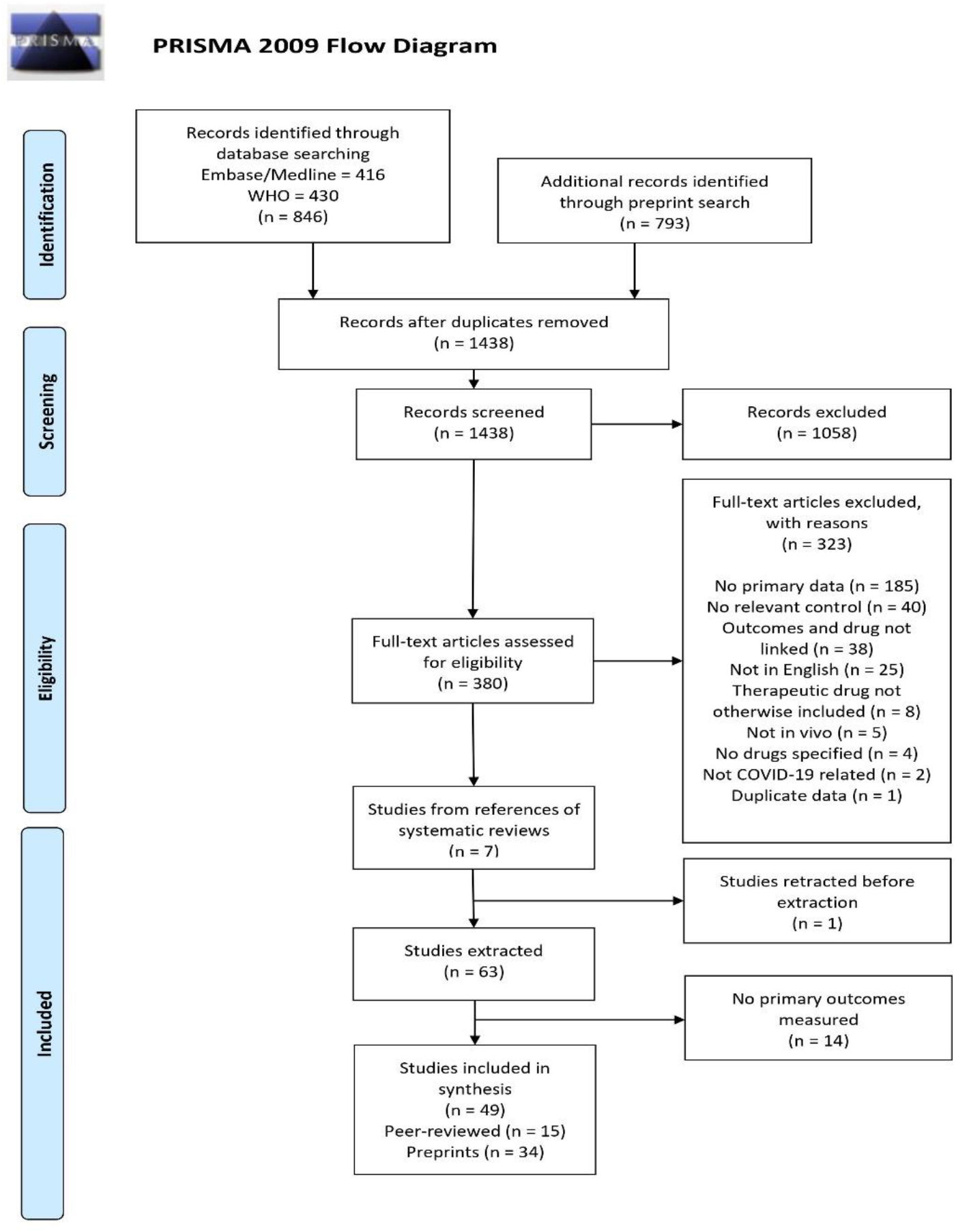
PRISMA Flow Chart. PRISMA - Preferred Reporting Items for Systematic Reviews and Meta-Analyses

### Study characteristics

51% of the papers identified were from China (n=25). The remainder were either from Europe (Denmark, France, Italy, Spain and United Kingdom (UK)), North America (United States of America (USA)) or other parts of Asia (South Korea) (Table 1). The majority of papers were cohort studies (n=29) and the remainder were either cross-sectional studies (n=9), case-control studies (n=8), case series (n=2) or a hybrid design (n=1). There were no experimental or quasi-experimental studies included in this review. Thirteen studies investigated more than one relevant drug, with one study (38) presenting data for all eight drug groups.

**Table 1.** Geographical Demographics of Included Studies and Peer-Review Status.

| Study Country | Number of studies (peer-reviewed) |
| --- | --- |
| China | 25 (10) |
| USA | 9 (2) |
| UK | 4 (0) |
| Italy | 3 (2) |
| Spain | 3 (1) |
| France | 2 (0) |
| South Korea | 2 (0) |
| Denmark | 1 (0) |
| Total | 49 (15) |

USA - United States of America, UK - United Kingdom.

### Exposure status and outcome measures

Most studies analysed outcomes for patients taking drugs regularly prior to contracting COVID-19. Studies that analysed ACEI, ARB, MCRA, statin and TZD usage measured outcomes only from those taking the drugs regularly, although one study compared outcomes of continued use of ACEIs and ARBs after admission with withdrawal of treatment (39). One study analysed acute treatment with the immunosuppressant tocilizumab (40), and one analysed acute treatment with the NSAID celecoxib (41). All the remaining studies, which looked at the use of drugs for treating COVID-19 focussed on corticosteroids. There was a mix of studies assessing acute treatment doses of corticosteroids and those taking long-term corticosteroids on severity of disease. All studies analysing susceptibility to infection with corticosteroids assessed those taking long-term steroids and all studies analysing mortality assessed those receiving acute doses.

### Quality assessment

The majority of the studies were assessed as being of fair quality (45%), followed by good quality (35%); the remaining studies were assessed to be of high risk of bias and rated as poor quality (20%). Generally, studies were downgraded due to limited justifications of sample sizes used and insufficient controlling for potential confounders.

#### Sensitivity analysis by publication status

As a considerable portion of the included studies were published as preprints that had not formally been peer-reviewed, we undertook sensitivity analysis on any of these studies eligible for meta-analysis to determine their influence on the overall effect estimates when included in random-effects models. None of the included preprints were found to significantly influence any of these results, described below, and they were therefore included in the analyses. Results of the sensitivity analyses can be viewed in Appendix 7.

### MCRAs, NSAIDs, Statins and TZDs

For four of the drug groups (MCRAs, NSAIDs, statins and TZDs), meta-analysis revealed no evidence of protection or harm for any of the groups of outcomes. None of these drug groups had any studies analysing mortality, and only NSAIDs had enough studies analysing severity of disease to perform a meta-analysis (Appendix 6 – Fig 14). No pooled effect estimates showed any statistically significant evidence of harm in patients with COVID-19 (Table 2) (Appendix 6 – Figs 15-17).

**Table 2.**
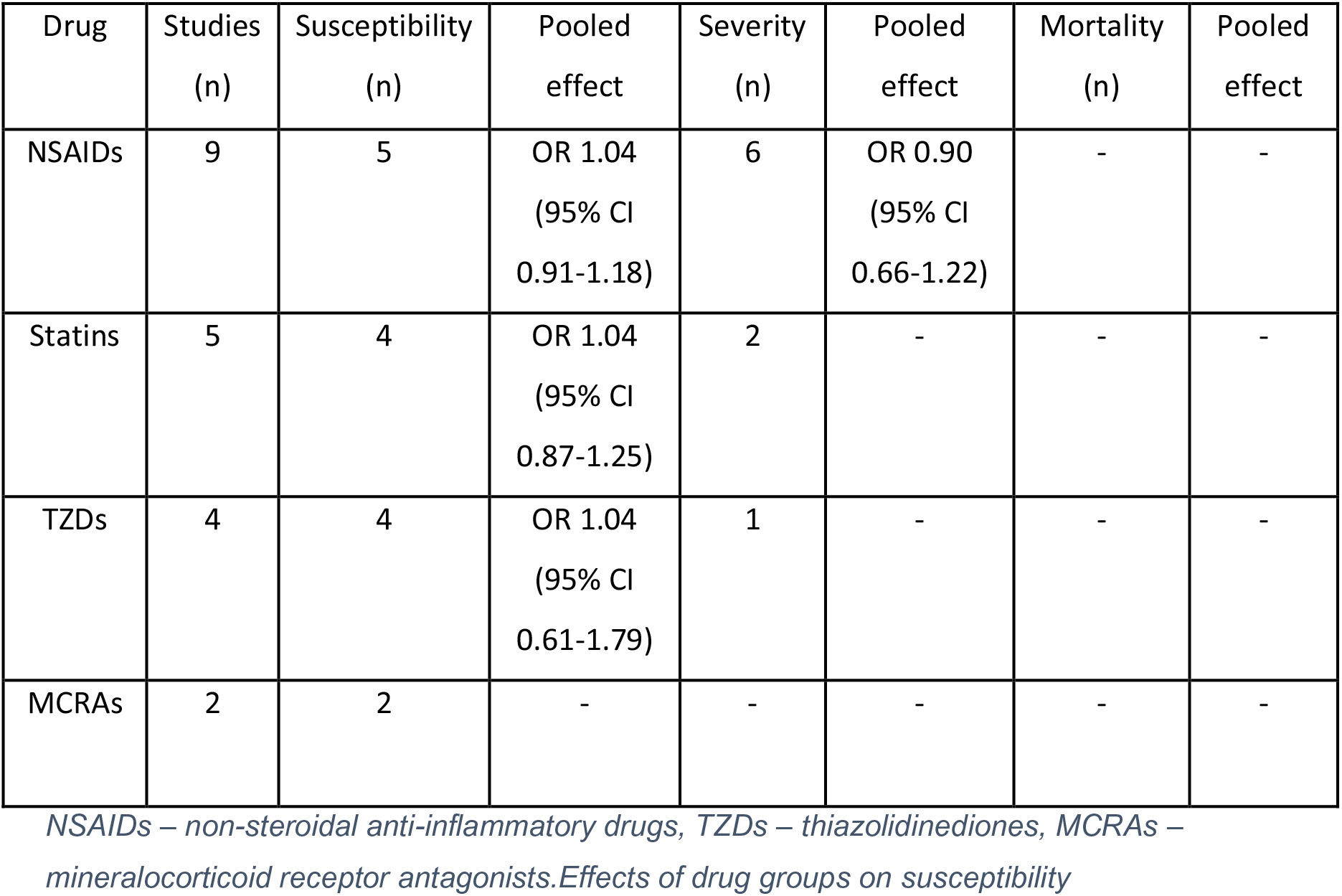
Summary of results for MCRAs, NSAIDs, statins and TZDs.

Results of studies looking into the effect of drug groups on the susceptibility to COVID-19 infection are summarised in Appendix 3 - **Table 3**.

#### RAASB

Ten studies were used in the meta-analysis of susceptibility to COVID-19 with RAASBs (33, 38, 42–49). Seven stratified their results into those taking ACEIs and those taking ARBs, and three combined the two drug groups (**Fig 2**).

**Fig 2.**
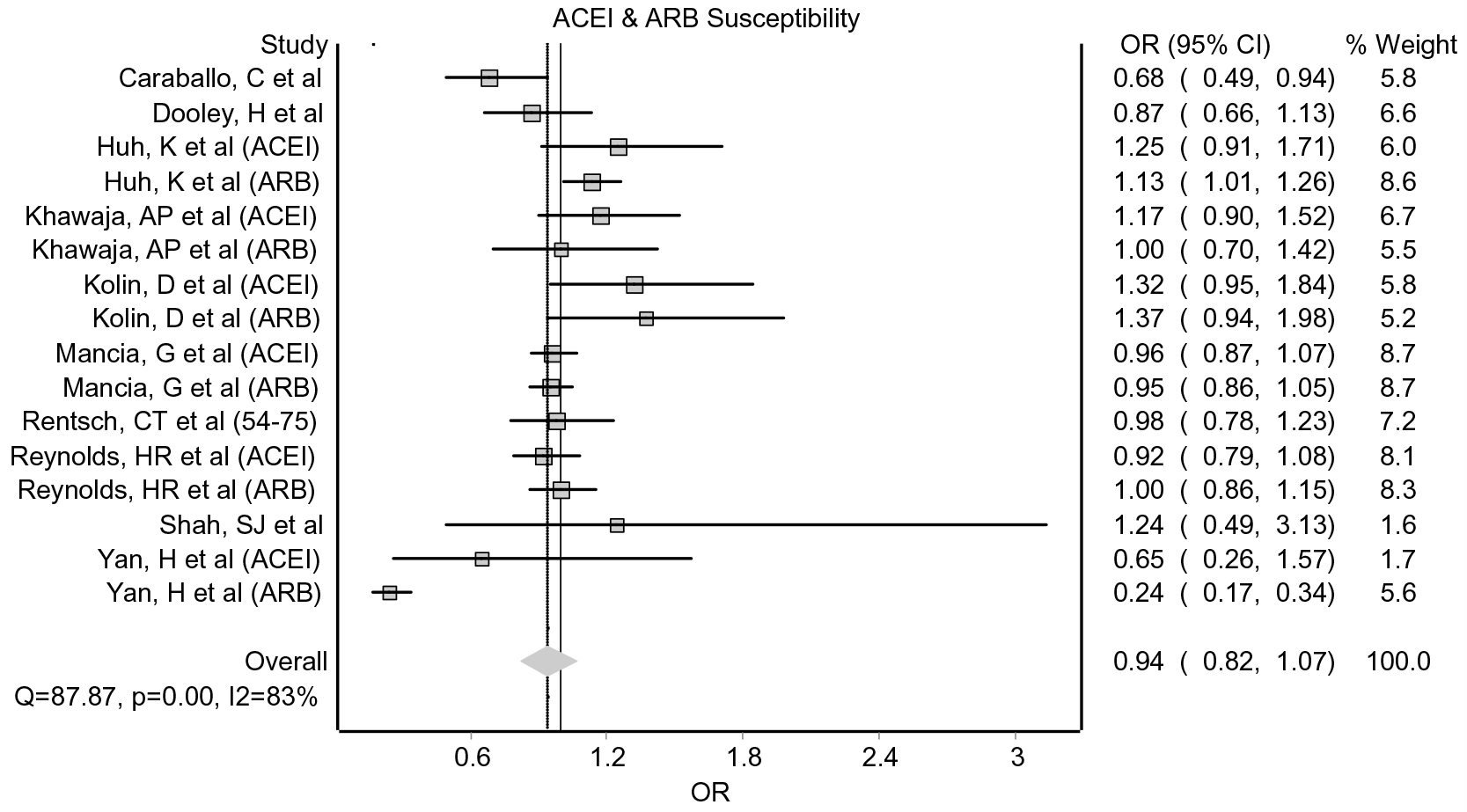
Forest plot for susceptibility to COVID-19 for those taking regular RAASBs. ACEI – angiotensin-converting enzyme inhibitor, ARB – angiotensin-II-receptor blocker, OR – odds ratio, CI – confidence interval.

The total pooled effect estimate from all the studies showed no evidence that RAASBs affect the risk of contracting COVID-19 (OR 0.94; 95% CI 0.82-1.07). Visual inspection of the funnel plot showed no evidence of publication bias (Appendix 4 - Fig 12). When split into subgroups for drug group, study design and statistical adjustment, this result was unchanged (Appendix 6 – Figs 18-20).

#### Corticosteroids

Four studies investigated the effects of corticosteroids on susceptibility to COVID-19 (38, 44, 48, 49). One study, rated as good quality, measured only those taking inhaled corticosteroids (38), therefore meta-analysis was not conducted. This study was also the only one to report an increased risk of contracting COVID-19 amongst patients taking corticosteroids, with the remaining three studies reporting null effects (Appendix 3 – **Table 3**).

#### Immunosuppressants

Five studies (32, 38, 44, 46, 48) analysing the effect of immunosuppressants on susceptibility to COVID-19 were included in the meta-analysis (**Fig 3**).

**Fig 3.**
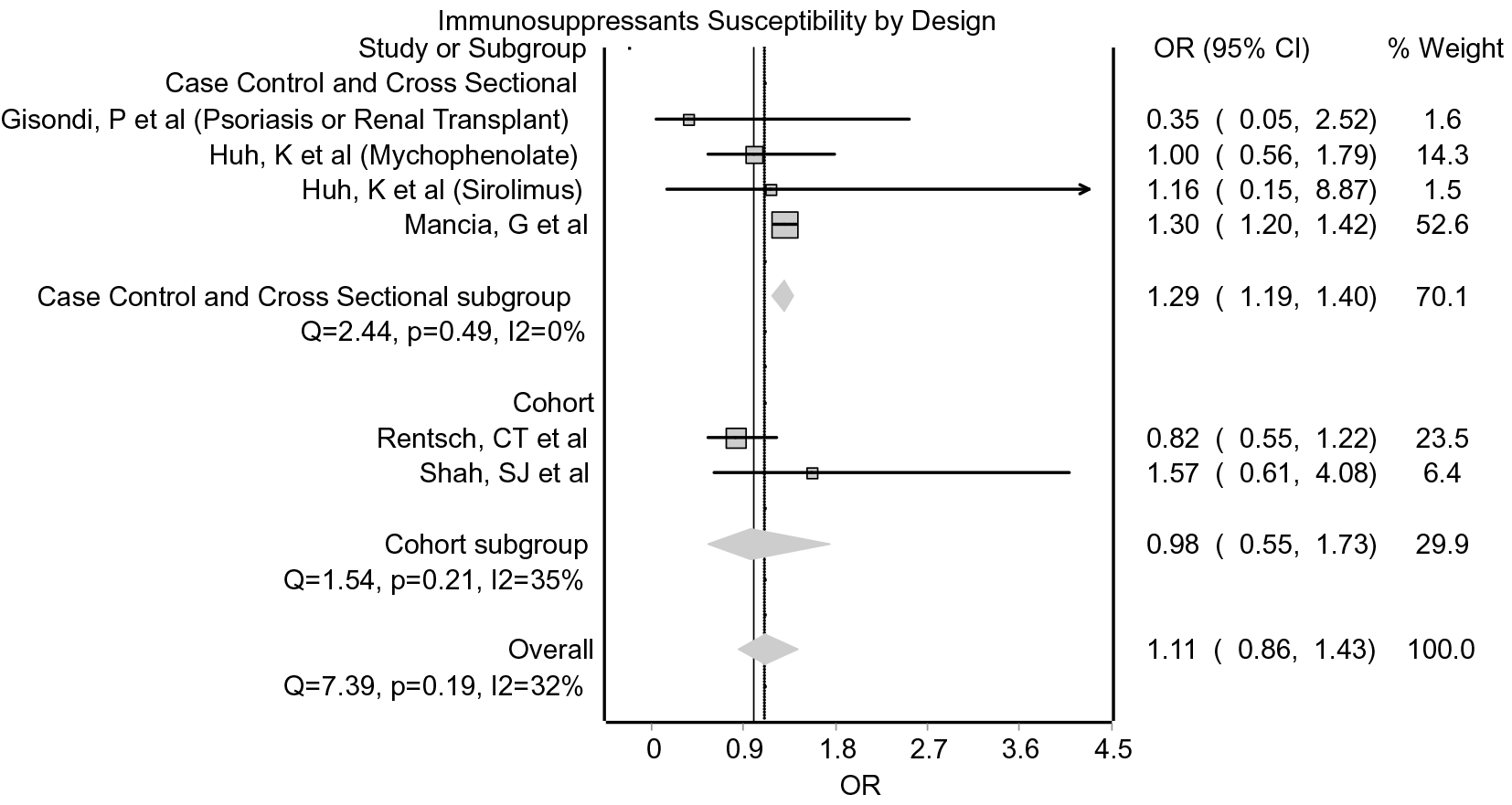
Forest plot for effect of immunosuppressants on susceptibility to COVID-19, stratified by study design. OR – odds ratio, CI – confidence interval.

The initial analysis showed no statistically significant evidence of harm or benefit (OR 1.11; 95% CI 0.86 – 1.43). Sensitivity analysis showed removal of Rentsch, CT et al resulted in a statistically significant movement of the pooled effect in the direction of harm (OR 1.29; 95% CI 1.19 - 1.40; I^2^ 0%), but was 96.9% weighted towards the result from Mancia, G et al. When analysed as subgroups based on study design, we found the same result for case-control and cross-sectional studies (OR 1.29; 95% CI 1.19 – 1.40) (**Fig 3**).

The only study looking at susceptibility that was not included in the meta-analysis (50) looked at individual immunosuppressive drugs compared against each other, finding janus kinase inhibitor usage was more prevalent among rheumatology patients with COVID-19, whereas no evidence of a protective or harmful effect of methotrexate was observed.

### Effects of drug groups on severity

Results of studies looking into the effect of drug groups on the severity of COVID-19 infection are summarised in Appendix 3 – **Table 4**.

#### RAASB

Among the total pool of studies, there was no evidence of benefit or harm with RAASBs (OR 0.93; 95% CI 0.77 – 1.12) (Fig 4). No individual studies affected either the pooled effect estimate or the heterogeneity. Visual inspection of the funnel plot showed no evidence of publication bias (Appendix 4 - Fig 13).

**Fig 4.**
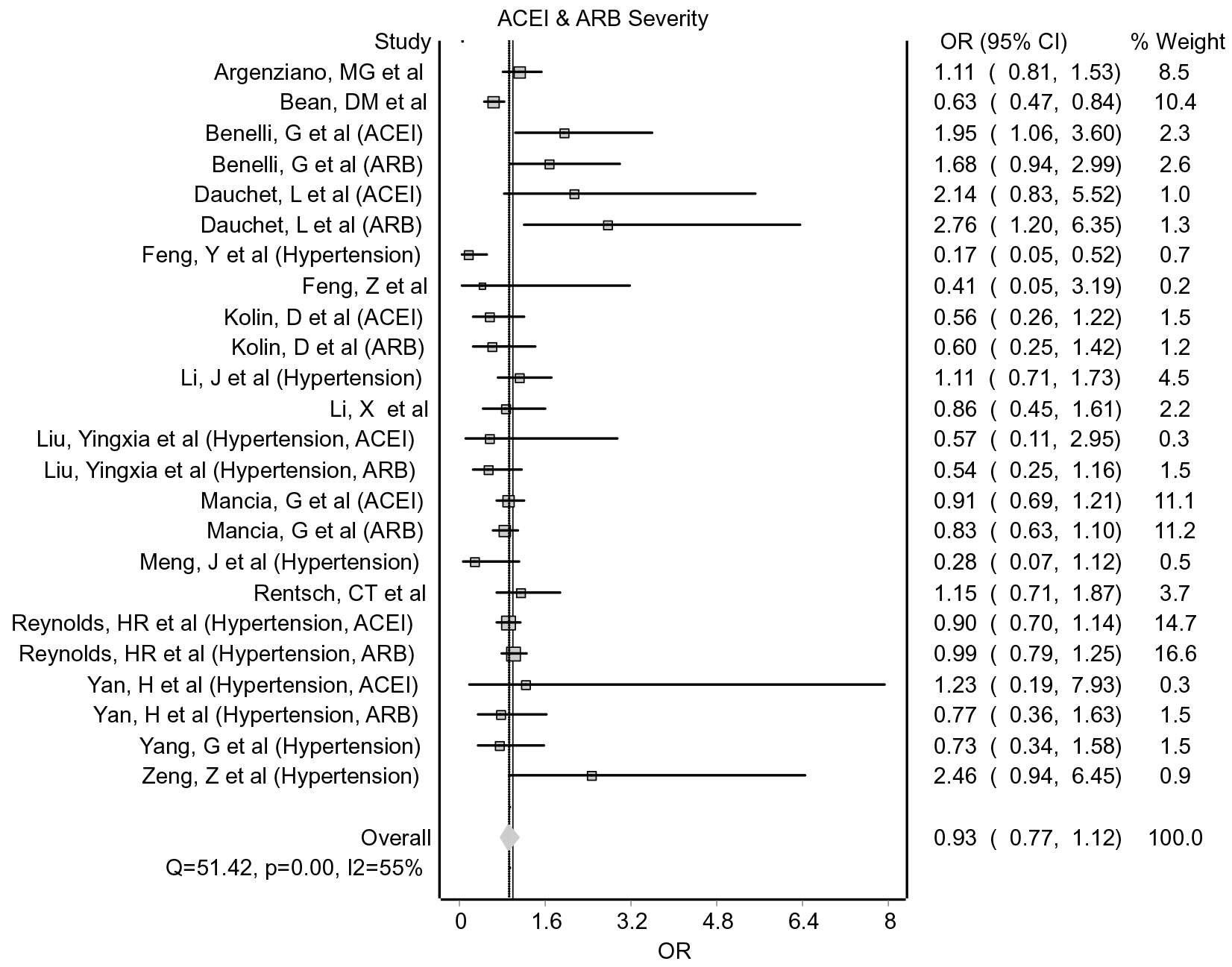
Forest plot of effect of RAASBs on severity of COVID-19. ACEI – angiotensin-converting enzyme inhibitor, ARB – angiotensin-II-receptor blocker, OR – odds ratio, CI – confidence interval.

Subgroup analyses consisting of three subgroups: either no or insufficient adjustment and compared against a general population; compared against hypertensive controls; and what we considered sufficiently adjusted results (adjustment for at least age, sex and one or more co-morbidities) was performed. This showed statistically significant evidence of a protective effect from ACEIs and ARBs for adjusted results (OR 0.86; 95% CI 0.77 – 0.96) (Fig 5). Sensitivity analysis showed that the removal of the result from Bean, DM et al (51) removed the statistically significant pooled effect estimate (OR 0.90; 95% CI 0.80 – 1.02).

**Fig 5.**
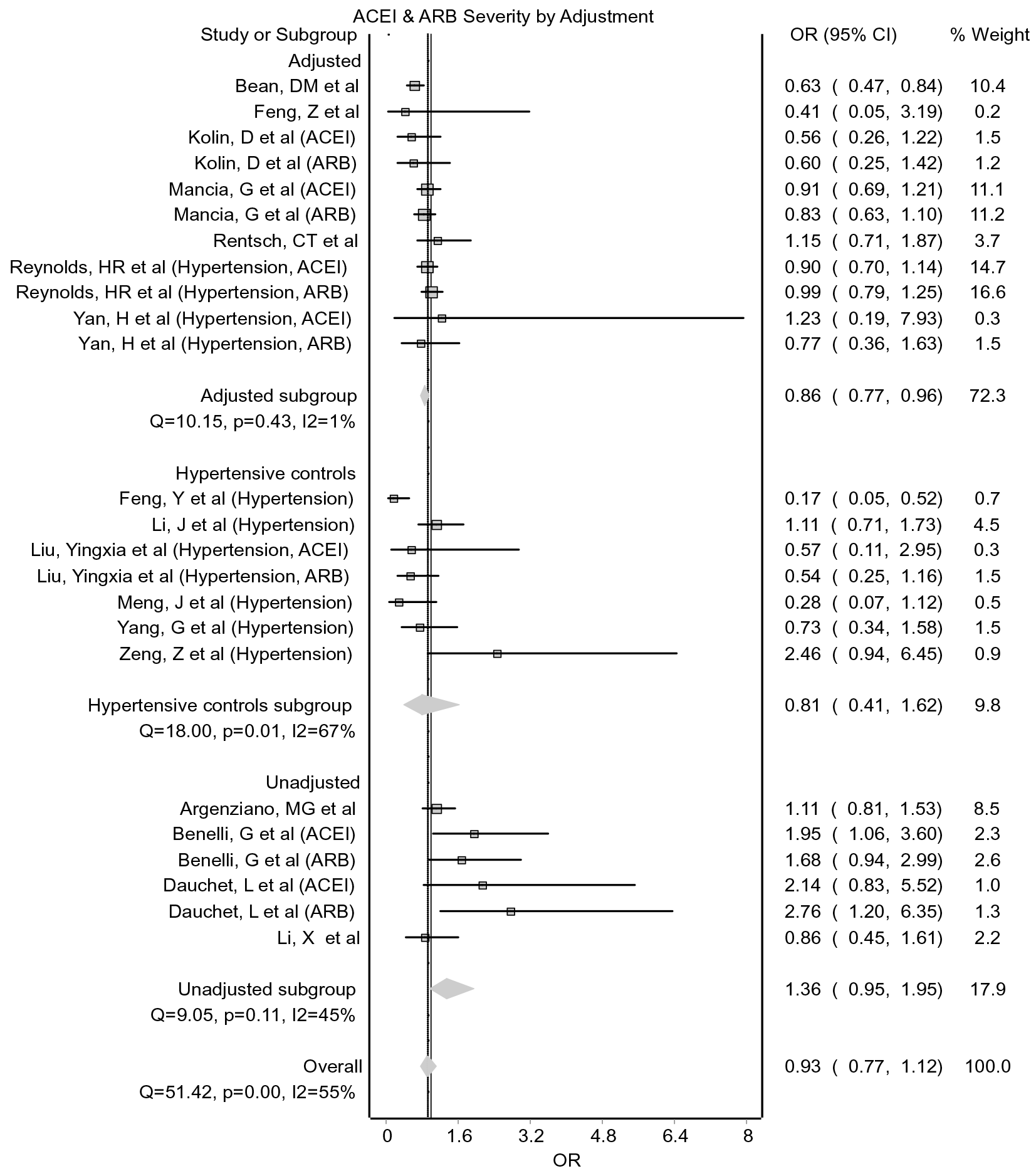
Forest plot of effect of RAASBs on severity of COVID-19 stratified by adjustment. ACEI – angiotensin-converting enzyme inhibitor, ARB – angiotensin-II-receptor blocker, OR – odds ratio, CI – confidence interval.

Creating diagnostic severity index or higher level of care subgroups showed statistically non-significant and heterogeneous results (OR 0.84; 95% CI 0.56 – 1.27; I^2^ 45% and OR 0.94; 95% CI 0.77 - 1.15; I^2^ 62%, respectively). When these two groups were analysed separately and subgroup analysis was run for level of adjustment, this showed a statistically non-significant result for adjusted studies using the severity index (OR 0.77; 95% CI 0.39 – 1.49). However, further statistically significant evidence of benefit for RAASBs among adjusted studies in regard to the highest level of care needed was observed (OR 0.85; 95% CI 0.74 – 0.98) (Fig 6). Again, removal of the result from Bean, DM et al (51) made the result statistically non-significant. This stratification also showed statistically significant evidence of harm from unadjusted studies (OR 1.63; 95% CI 1.16 - 2.31).

**Fig 6.**
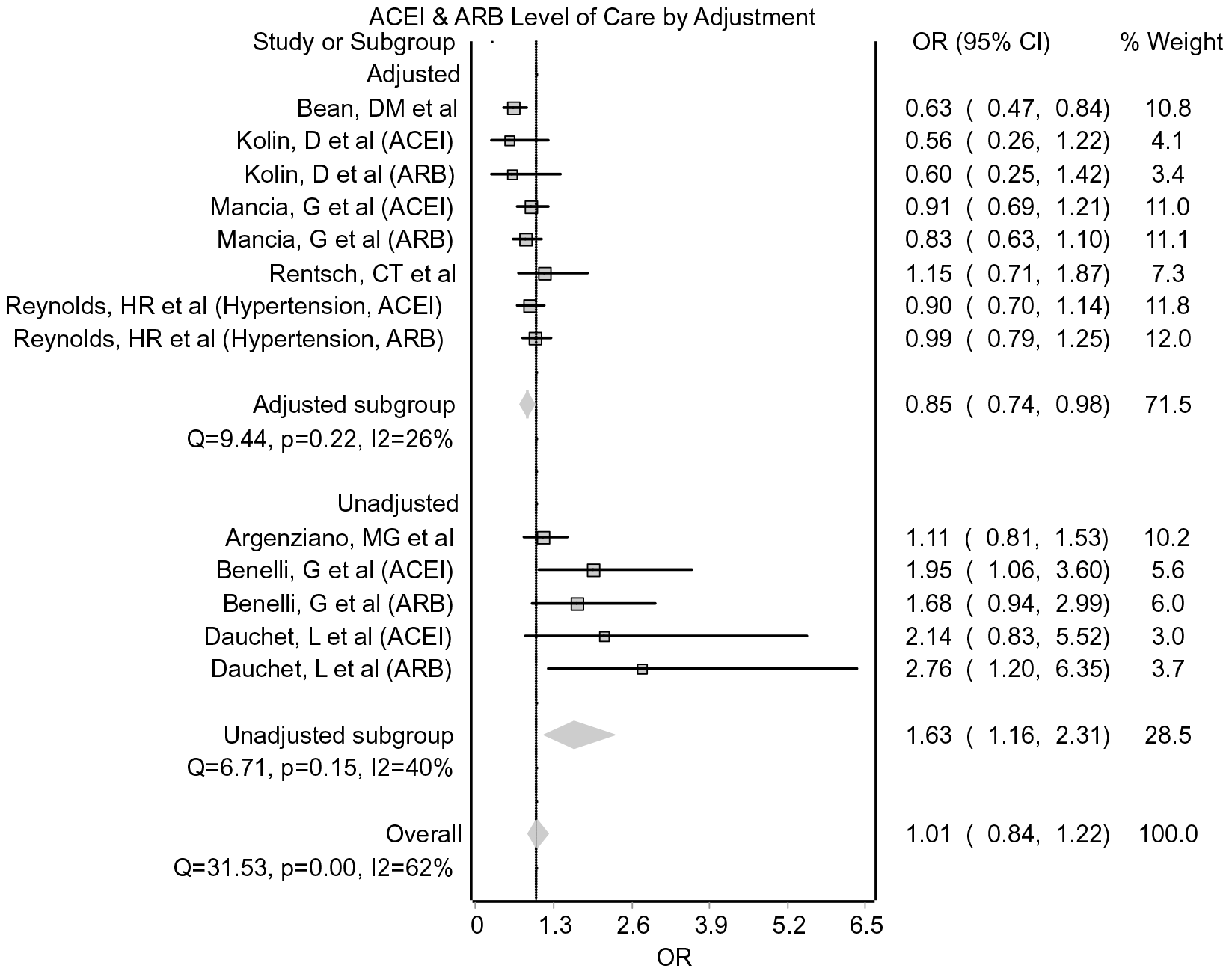
Forest plot of effect of RAASBs on the highest level of care needed, stratified by level of adjustment. ACEI – angiotensin-converting enzyme inhibitor, ARB – angiotensin-II-receptor blocker, OR – odds ratio, CI – confidence interval.

#### Corticosteroids

Eight studies with verifiable data assessed the risk of corticosteroids increasing the risk of developing severe disease (49, 52–58). The majority were cohort studies, which varied by study quality and other methodological factors (Appendix 3 – **Table 4**).

Four cohort studies measured outcomes from acute treatment doses of corticosteroids, reaching different conclusions; however, one study did not have a calculable OR and so a meta-analysis was not possible.

The four studies which used long-term steroids as their exposure were analysed together in a meta-analysis. One study ran different analyses for oral and inhaled steroids. As these can be co-prescribed, this analysis was run twice with each model including either the inhaled or oral corticosteroid measurement. Neither model showed statistically significant evidence of harm or benefit for severity of COVID-19 for those taking long-term steroids (inhaled corticosteroids = OR 1.40; 95% CI 0.81 – 2.39; I^2^ 36%; oral corticosteroids = OR 1.36; 95% CI 0.76 – 2.45; I^2^ 44% (**Fig 7**)).

**Fig 7.**
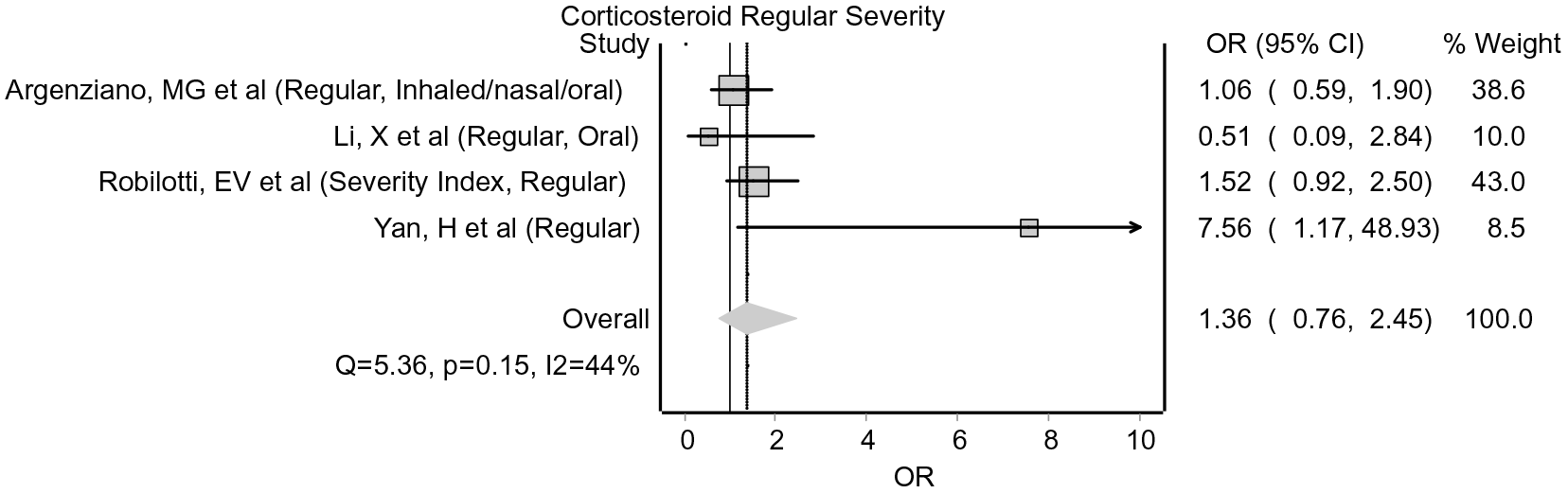
Forest plot of effect of regular corticosteroids on severity of COVID-19. OR – odds ratio, CI – confidence interval.

#### Immunosuppressants

Meta-analysis was conducted on four studies (46, 54, 56, 59) of severity of COVID-19 in those taking immunosuppressants. All four were cohort studies and all scored poorly on aspects of controlling for confounding during quality assessment (Appendix 3 – **Table 4**).

The pooled effect estimate showed no statistically significant evidence of benefit or harm (OR 0.68; 0.27 – 1.71) (**Fig 8**).

**Fig 8.**
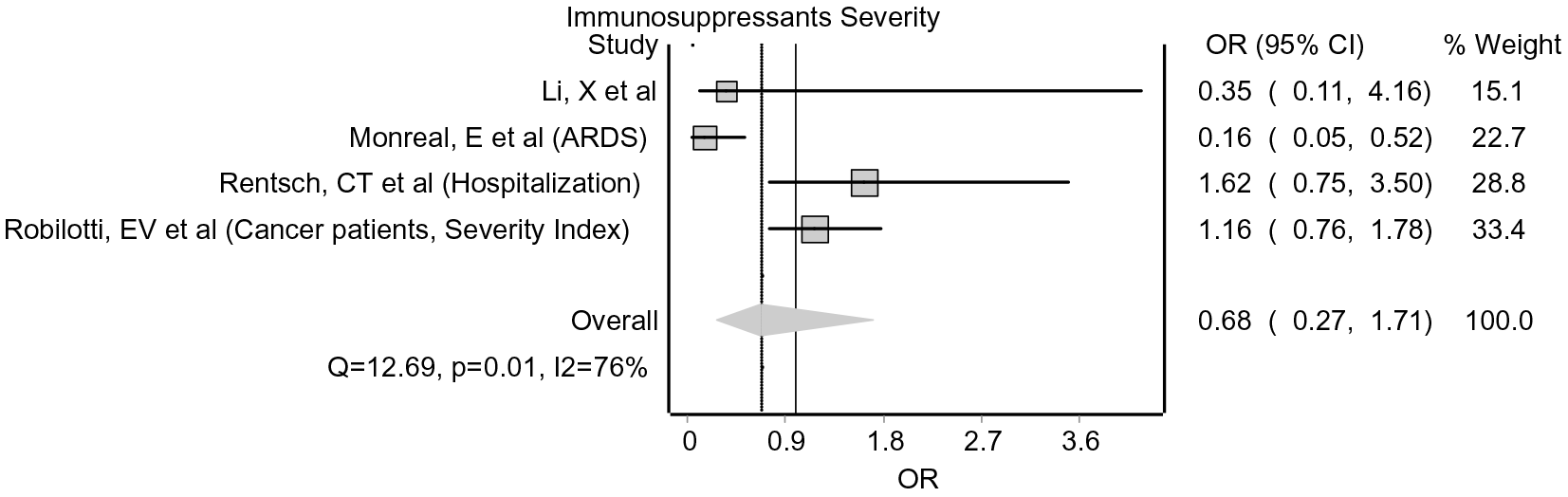
Forest plot for effect of immunosuppressants on severity of COVID-19. OR – odds ratio, CI – confidence interval, ARDS – acute respiratory distress syndrome.

The studies not included in this meta-analysis reached opposing conclusions, but in very different scenarios. The only study to look at acute usage of tocilizumab (40) was a case control study, finding evidence of benefit in terms of requiring ventilation (OR 0.42: 95% CI 0.2 – 0.89) or ICU care (OR 0.17; 95% CI 0.06-0.48). An additional case control study (60), found that those on immunosuppressants had more severe COVID-19 when compared with their family members who tested positive for SARS-CoV-2, but no OR was available nor calculable.

### Effects of drug groups on mortality

Results of studies looking into the effect of drug groups on mortality in COVID-19 infection are summarised in Appendix 3 – **Table 5**.

#### RAASB

Eight studies investigated death rates among those taking RAASBs (61–68) with six grouping them together and two analysing ACEIs and ARBs separately (Appendix 3 – **Table 5**).

None of these studies adjusted their results, although three used a hypertensive control group. When subgroup analysis was undertaken to reflect these, it showed no evidence of harm or benefit against hypertensive controls (OR 0.91; 95% CI 0.62 – 1.32), the general population (OR 1.11; 95% CI 0.67 – 1.85) or for the pooled effect overall (OR 0.97; 95% CI 0.74 – 1.29) (Fig 9).

**Fig 9.**
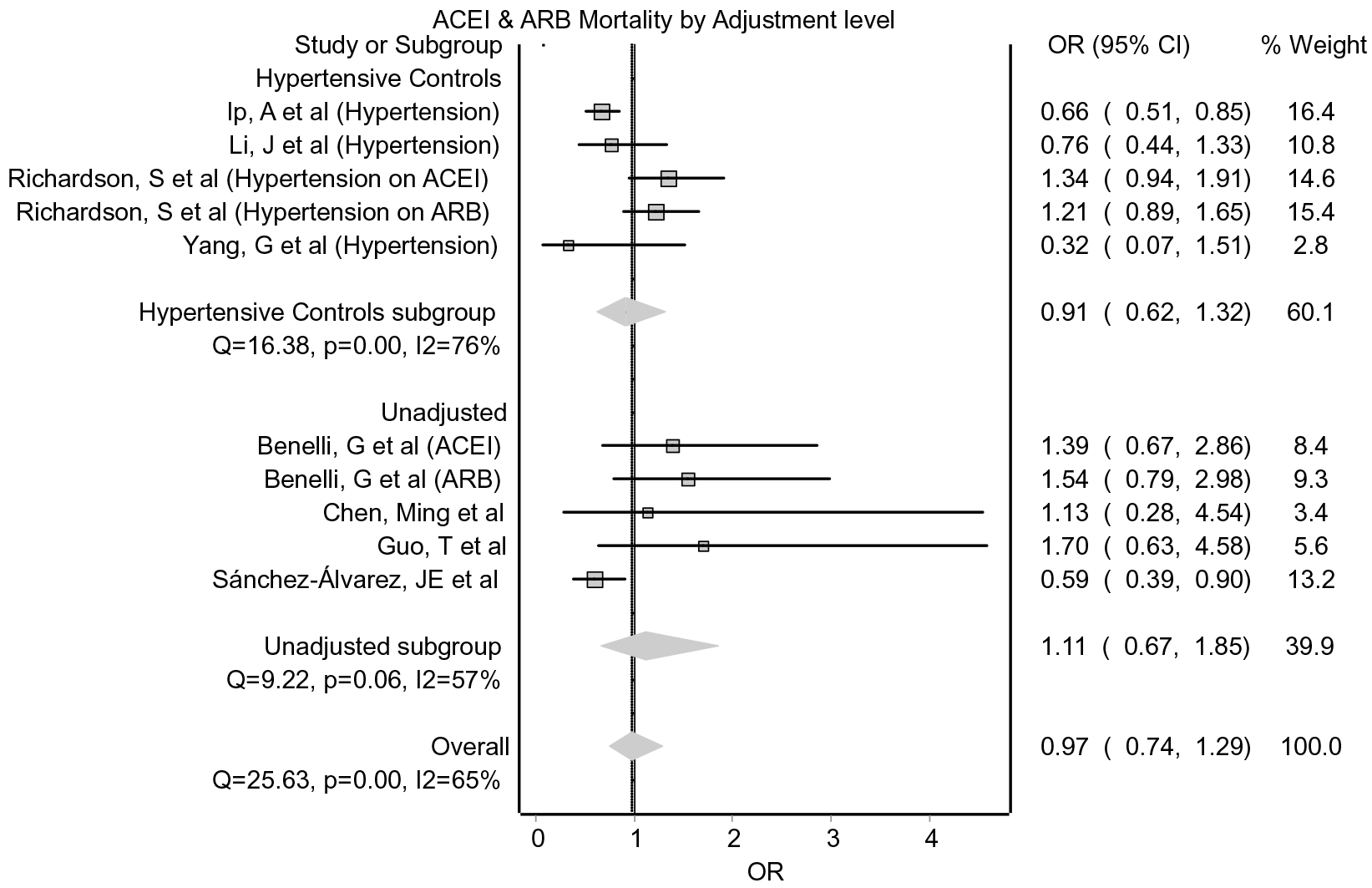
Forest plot of effect of RAASBs on mortality, stratified by control group. ACEI – angiotensin-converting enzyme inhibitor, ARB – angiotensin-II-receptor blocker, OR – odds ratio, CI – confidence interval.

Subgroup analysis of studies which combined RAASBs showed a pooled effect estimate exhibiting statistically significant evidence of benefit (OR 0.68; 95% CI 0.55 – 0.85) with low heterogeneity (I^2^ 5%) (Fig 10). During sensitivity analysis, the result became statistically non-significant if the results from either Ip, A et al or Sánchez-Álvarez, JE et al were removed. These two studies were also the only two in this analysis that were rated as poor during the quality assessment.

**Fig 10.**
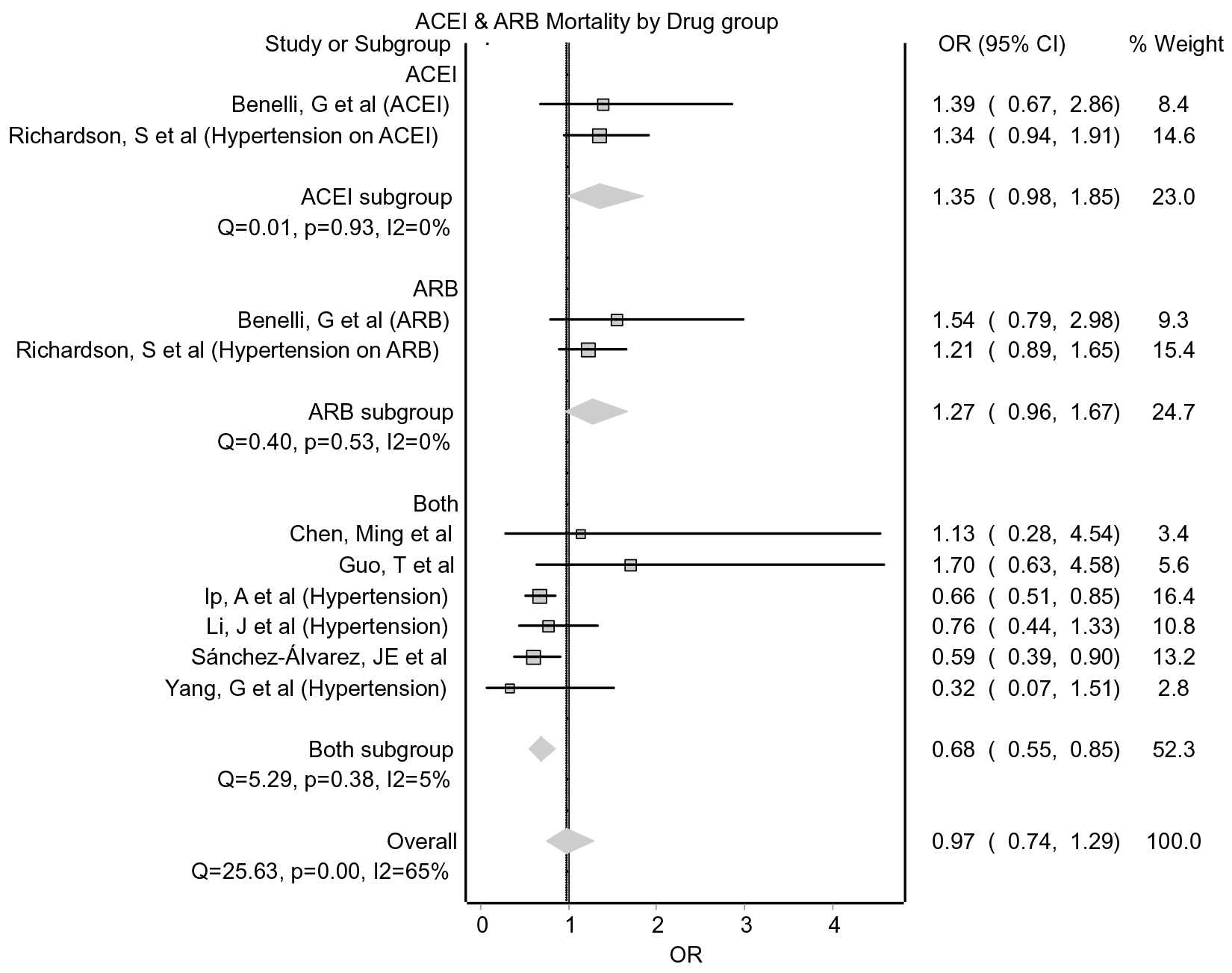
Forest plot for effect of RAASBs on mortality in COVID-19, stratified by drug group. ACEI – angiotensin-converting enzyme inhibitor, ARB – angiotensin-II-receptor blocker, OR – odds ratio, CI – confidence interval.

Two studies were not included in the meta-analysis, each reporting conflicting results with regards to mortality (69, 70) (Appendix 3 – **Table 5**).

#### Corticosteroids

Seven (62, 67, 71–75) of the 10 studies assessing mortality in those given corticosteroids were included in the meta-analysis. All of these analysed acute doses used to treat COVID-19 admissions in hospital (Appendix 3 – **Table 5**). Two studies (55, 76) reported no evidence of harm or benefit but it was not possible to extract summary data for pooled analysis. The remaining study (54) compared hazard ratios (HRs) for high and low dose corticosteroids, finding a low-dose had no evidence of either harm or benefit (HR 1.26; 95% CI 0.61 – 2.58), but a high-dose showed evidence of increased mortality (HR 3.5; 95% CI 1.79 – 6.86).

Meta-analysis of the remaining studies showed evidence of harm (OR 2.22; 95% 1.26 – 3.90) (Fig 11). The authors of three of the studies highlighted confounding by indication as a likely factor considerably affecting their results, and when these studies were removed, the pooled effect estimate showed there was no clear evidence of an effect (OR 1.48; 95% CI 0.73 – 3.00; I^2^ 67%).

**Fig 11.**
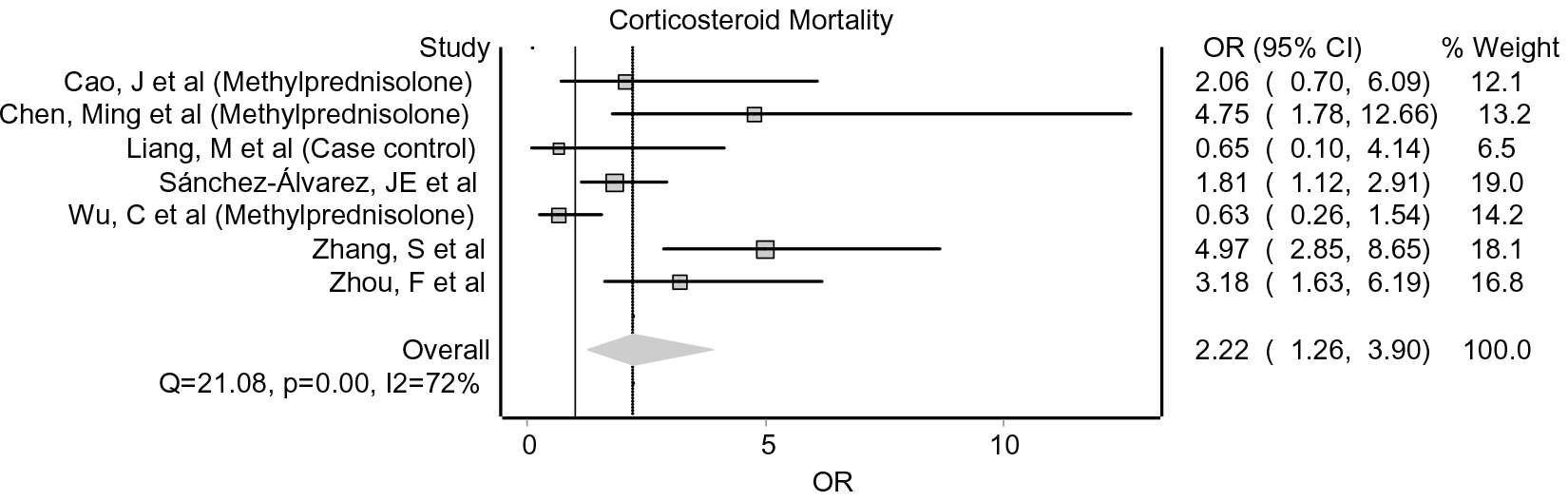
Forest plot of effect of corticosteroids on mortality. OR – odds ratio, CI – confidence interval.

#### Immunosuppressants

Two studies analysed mortality rates in those taking immunosuppressants. (40, 59). Neither found statistically significant evidence of increased or decreased mortality (OR 0.88; 95% CI 0.17 – 4.46 and OR 0.90; 95% CI 0.24 – 3.42, respectively) (Appendix 3 – **Table 5**).

## Discussion

### Statement of principal findings

This rapid systematic review and meta-analysis attempted to identify and assess any potentially deleterious drug groups in covid-19 susceptibility and prognosis. In terms of susceptibility to infection, we found no pooled effect estimates and one subgroup analysis with statistically significant evidence of increased susceptibility (Fig 3), which would be in keeping with many other infections and a known risk of immunosuppressant use (77). Hence, we found no evidence to suggest that those without COVID-19 should stop taking their medication to reduce their risk of contracting the disease. Stopping immunosuppressants may also result in a flare-up of a person’s underlying condition, which can result in increased risk of infection (78). Furthermore, the withdrawal of any necessary medications could cause harm from the disease the drug was being used to treat, as well as from a potentially severe form of COVID-19 for people with hypertension and diabetes who are already at increased risk (79, 80).

We found no evidence that long-term use of any of these drugs increases the severity of disease, nor the mortality rates. One subgroup analysis of RAASBs showed a statistically significant relationship between RAASB usage and requiring a higher level of hospital care (**Fig 6**). However, this was found in an unadjusted subgroup where the adjusted subgroup showed evidence of protection, highlighting the effect of known confounders, such as age and cardiovascular disease (81), on the results from these observational studies. This evidence of protection against more severe disease was also shown by the pooled effect of all adjusted studies analysing severity (Fig 5).

### Strengths and limitations

This rapid systematic review and meta-analysis has synthesised the best available evidence from the first few months of the pandemic for each of these drugs when taken by those with or at risk of COVID-19. This study has, within the limitations imposed by time and the unparalleled research publication rate, aimed to compile a comprehensive list of drugs that were hypothesized to be deleterious in peer-reviewed papers at the time of the searches.

This review however has several limitations. The rapidity with which research is being produced and published affected our study in a number of ways. Firstly, we acknowledge that while a rapid review is justifiable on the grounds of swifter completion, it is less robust than a systematic review. Secondly, the studies included in this review were also completed within a short space of time in order to attempt to learn from clinical practice as quickly as possible. As a result of this, many papers that would be relevant to this review, including papers with stronger evidence and more robust methodology than some of those included may now be available. Finally, the research landscape is changing so quickly that the studies included in this review were all published prior to conducting the last search. This will bias our results towards the studies that were produced more quickly and in countries that experienced larger COVID-19 case numbers earlier on in this outbreak. These countries may have large differences in their demographics as well as clinical protocols for both pre-existing co-morbidities and COVID-19, which could impact the generalisability of our findings.

Another factor heightening the risk of bias, errors and methodological inadequacies, is the lack of peer-reviewed articles and the use of preprints in this review. We conducted a sensitivity analysis in January 2021 to analyse whether the removal of papers that had not made it through the peer-review process at this point affected the results, but there were no significant changes to the pooled outcomes (Appendix 7). Nevertheless, we must emphasise that because we have included preprints, the results from this review are only to highlight potential outcomes to evaluate with further studies and should not be used to alter clinical practice.

As all of the evidence in this review is from observational studies, this introduces a risk of bias from confounding factors, despite many studies attempting to statistically adjust for these. Any confounders not adjusted for or as yet unknown will have affected our results. One example of this is the discovery that people with a Black, Asian or minority ethnic background are at an increased risk of severe COVID-19 (82), something that was not apparent at the beginning of the outbreak and was therefore only adjusted for in five of the studies included in the review. Other biases evident in the studies included confounding by indication, most evident among the studies looking at acute treatment of COVID-19 with corticosteroids, and misclassification bias in studies where it was possible the control group had been buying the drug of interest over-the-counter or had stopped taking it immediately prior to admission to hospital. This happened with NSAIDS for example.

Using aspects of the GRADE assessment, the quality of evidence presented in this review could be classified as low, primarily due to inconsistency, indirectness, imprecision and risk of bias across each outcome (31).

### Interpretation in light of the wider literature

One of the reasons RAASBs, MCRAs, NSAIDs, Statins and TZDs were hypothesized to be harmful is their potentially upregulating effect on ACE2, this being the entry point into cells for SARS-CoV-2. Although there are studies which have shown that all of these drugs may upregulate ACE2, particularly RAASBs, the evidence is largely from animal studies, and some of it is contradictory (83). We found no evidence to suggest any of these drugs worsen outcomes or increase the susceptibility to contracting COVID-19 and some evidence to suggest RAASBs could be protective. Whilst it is unclear if this is related to their effect on ACE2, it appears that the protective nature of ACE2 may in fact provide a target for treating COVID-19 (84). Studies have also shown similar protective effects of ACEIs (85), ARBs and statins (86) in non-COVID pneumonias.

Another prominent reason for drugs to be hypothesized to worsen outcomes was immunosuppression. Advice emerged early on advising against using steroids in COVID-19 (14), but this was at odds with the experiences of front-line clinical workers in China who were using it in a large number of cases (87). Guidance surrounding long-term immunosuppressants in those with COVID-19 was also issued (88), however at the same time it appeared that dampening the immune response may have a role in treating acute infections (89). With the discovery that interleukin 6 (IL-6) was elevated in patients with severe COVID-19 (90), IL-6 antagonists such as tocilizumab started to be used therapeutically (91). We only found one study which analysed the outcomes of those treated with tocilizumab and this found a reduction of ICU admissions and need for ventilation when patients were given tocilizumab (40). This was an observational study where the patients were “highly selected” so, although the results were adjusted for confounders, there remains a risk of bias.

The majority of studies included in this review measured outcomes in patients taking drugs regularly. We only found one study (39) comparing patients who had the drug withheld during the acute infection with those that continued to take them. This study found no effect on disease severity or mortality but found statistically significant reduction in viral clearance and length of stay in those in whom RAASBs were withheld during their hospital stay. This area requires more research, as any conclusions we draw from this review can only be applied to those who take these medications prior to the onset of COVID-19. In order to guide clinical practice, studies comparing initiated, continued and withheld medications, ideally randomised controlled trials (RCTs), are paramount (92).

With most drug groups included in this review, dosage and timing was not usually measured or analysed. One study of corticosteroids compared the effect of the cumulative dose of steroids and the authors concluded that mortality with high-dose steroids was higher than with low-dose steroids (54). Another study which found favourable outcomes for corticosteroids as a treatment for COVID-19 hypothesized that the timing was also important, opting for low dose steroids early on in the disease progression (93). These hypotheses were similarly at risk of confounding by indication, with those going on to develop ARDS more likely to get higher dose steroids at a later point in time.

### Implications for policy, practice and research

This review contains exclusively observational studies, many of which have not been peer-reviewed, and so we do not recommend any changes to normal prescribing practice. However, we have found no evidence to suggest medications should be stopped solely due to contracting COVID-19. This goes along with guidelines published for many of these drug groups (94–96) and the findings of other rapid reviews (28, 97).

For those taking the drugs highlighted in this review, the binary decision between stopping and continuing their medications, coupled with the contrasting advice from experts, the media and governments, helped fuel the false dichotomy that drug groups were either harmful or not. Due to the large number of factors that can affect outcomes in COVID-19, the harm or benefit derived from starting, stopping or continuing drugs will affect individuals differently, with co-morbidities, demographics, other medications and individual response to the virus playing a role in the progression of the disease. However, this review raises the prospect that drug groups that have been hypothesized to be deleterious in COVID-19, may have the potential to be beneficial in certain circumstances. This is evidenced further by a number of clinical trials, either planned, in progress or completed, involving drugs or drug groups identified during this review (98–103).

Furthermore, where trials are unfeasible, as in the case where the aim is to examine the effects of long-term drug use on COVID-19, good quality, large-scale observational studies will be necessary to form the best possible evidence. RCTs will understandably be focussed on finding drugs used to treat COVID-19, the results of which are beginning to be published (98). However, trials looking at treatments with drugs included in this review may help inform their use in other contexts.

## Conclusions

We have highlighted the gaps in the research, such as the lack of RCTs and mortality data for NSAIDs, immunosuppressants and long-term steroid use. We have proposed that there should be some focus in future research on these gaps as well as the use of ACE2, or drugs which upregulate it, as a potential target for treating COVID-19.

This study found a total of eight drug groups hypothesized to be deleterious in COVID-19. The available data from the first few months of the current pandemic suggest that there is little to no evidence these drug groups increase susceptibility, severity or mortality in COVID-19 and we found some evidence that ACEIs and ARBs may be protective in preventing a more severe disease.

## Data Availability

All data publicly available through papers included in analysis

## Appendix 1: search terms

### First search (Ovid)

1. Coronavirus Infections/ or covid-19.mp. or Betacoronavirus/
2. Pneumonia, Viral/ or Severe Acute Respiratory Syndrome/ or sars-cov-2.mp. or SARS Virus/
3. Pandemics/ or ncov-2019.mp.
4. 2019-nCov.mp. or Spike Glycoprotein, Coronavirus/
5. 1 or 2 or 3 or 4
6. Analgesics/ or Anti-Inflammatory Agents, Non-Steroidal/ or non-steroidal.mp or Cyclooxygenase 2 Inhibitors/
7. Ibuprofen.mp. or ibuprofen/
8. Angiotensin-Converting Enzyme Inhibitor?.mp. or Angiotensin-Converting Enzyme Inhibitors/
9. ACE Inhibitor?.mp.
10. Antihypertensive Agents/ or Angiotensin Receptor Antagonists/ or ARB?.mp. or Angiotensin II Type 1 Receptor Blockers/
11. Angiotensin Receptor Blocker?.mp.
12. Angiotensin II Receptor Blocker?.mp.
13. Steroid?.mp. or Steroids/
14. Corticosteroid?.mp. or Adrenal Cortex Hormones/
15. Immunosuppressant?.mp. or Immunosuppressive Agents/
16. ((drug? or medication? or prescri*) adj10 (harm or safe* or risk or worse* or sever* or discontinue? or unsafe or avoid)).mp. [mp=ti, ab, ot, nm, hw, fx, kf, ox, px, rx, ui, an, sy, tn, dm, mf, dv, kw, dq]
17. 6 or 7 or 8 or 9 or 10 or 11 or 12 or 13 or 14 or 15 or 16
18. 5 and 17
19. Limit 18 to yr=”2019-Current”

### Second search (WHO COVID-19 database)

1. NSAID*
2. Non-steroid*
3. Analgesi*
4. Ibuprofen
5. ACE Inhibitor*
6. ACE-I*
7. ACEI*
8. Angiotensin Converting Enzyme Inhibitor*
9. Renin-Angiotensin-Aldosterone System Inhibitor*
10. Renin-Angiotensin-Aldosterone System Blocker*
11. RAAS Inhibitor*
12. RAAS Blocker*
13. *pril
14. Antihypertensive*
15. Angiotensin Receptor Blocker*
16. ARB*
17. *sartan
18. *steroid*
19. *cortico*
20. Immunosuppressant*
21. Immunosuppressive*
22. Immunomodulat*
23. Immune modulat*
24. Antimineralocorticoid*
25. Aldosterone antagonist*
26. Spironolactone
27. MCRA*
28. MRA*
29. Mineralocorticoid Receptor Antagonist*
30. *statin*
31. HMG CoA Reductase Inhibitor*
32. Thiazolidinedione*
33. *glitazone*

### Third search (NIH iSearch COVID-19 portfolio)

1. NSAID
2. Non-steroid
3. Analgesi*
4. Ibuprofen
5. ACE Inhibitor
6. ACE-I
7. ACEI
8. Angiotensin Converting Enzyme Inhibitor
9. “Renin-Angiotensin-Aldosterone System Inhibitor”
10. “Renin-Angiotensin-Aldosterone System Blocker”
11. “RAAS Inhibitor”
12. “RAAS Blocker”
13. *pril
14. Antihypertensive
15. Angiotensin Receptor Blocker
16. ARB
17. *sartan
18. *steroid*
19. *cortico*
20. Immunosuppressant
21. Immunosuppressive
22. Immunomodulat*
23. “Immune modulator”
24. Antimineralocorticoid
25. “Aldosterone antagonist”
26. Spironolactone
27. MCRA
28. MRA
29. “Mineralocorticoid Receptor Antagonist”
30. *statin
31. “HMG CoA Reductase Inhibitor”
32. Thiazolidinedione
33. *glitazone

## Appendix 2: inclusion & exclusion criteria

### First set

Inclusion criteria:

1. Papers looking at whether a medication or medications are potentially unsafe if prescribed for patients with confirmed or suspected COVID-19 or SARS-Cov-2 infection
2. Papers looking at whether a medication or medications should be discontinued in patients who have confirmed or suspected COVID-19 or SARS-Cov-2 infection and would otherwise take them regularly
3. Papers looking at the overall safety or benefits of prescribing or discontinuing a medication or medications that have been identified as potentially unsafe for patients under criteria 1 or 2
4. Paper addressing or responding to any paper already included in reference to the safety of a medication or medications

Exclusion criteria:

1. Paper does not mention any specific medications or groups of medications
2. Paper not related to current COVID-19 outbreak
3. Published before November 2019
4. Paper not written in English
5. Only looking at unlicensed uses of medications, where no medication involved has already been identified as posing a potential risk when prescribed for its licensed use(s)
6. Only looking at medications being used for the treatment of COVID-19, where no medication involved has already been identified as posing a potential risk when prescribed for its licensed use(s)
7. No new data, hypothesis, guidance or recommendations stated related to COVID-19 for any individual relevant medications

### Second set

Inclusion criteria:

1. Studies looking at the outcomes of patients with COVID-19 or SARS-Cov-2 infection who were, or would normally be, taking medications that have been identified in the preliminary search as potentially harmful if prescribed for patients with confirmed or suspected COVID-19 or SARS-Cov-2 infection (see list)
2. Studies looking at the outcomes of patients with COVID-19 or SARS-Cov-2 infection when given medications that have been identified in the preliminary search as potentially harmful if prescribed for patients with confirmed or suspected COVID-19 or SARS-Cov-2 infection

Exclusion criteria:

1. Paper does not mention any medications or groups of medications already identified as potentially harmful
2. Paper not related to current COVID-19 outbreak
3. Published before November 2019
4. Paper not written in English
5. Paper has no primary data
6. Study not performed in vivo
7. It is unclear from the paper if the clinical outcomes follow the administration of a medication or group of medications already identified as potentially harmful
8. There is no measure against non-COVID-19 or non-drug exposed control

## Appendix 3: study tables

**Table 3.**
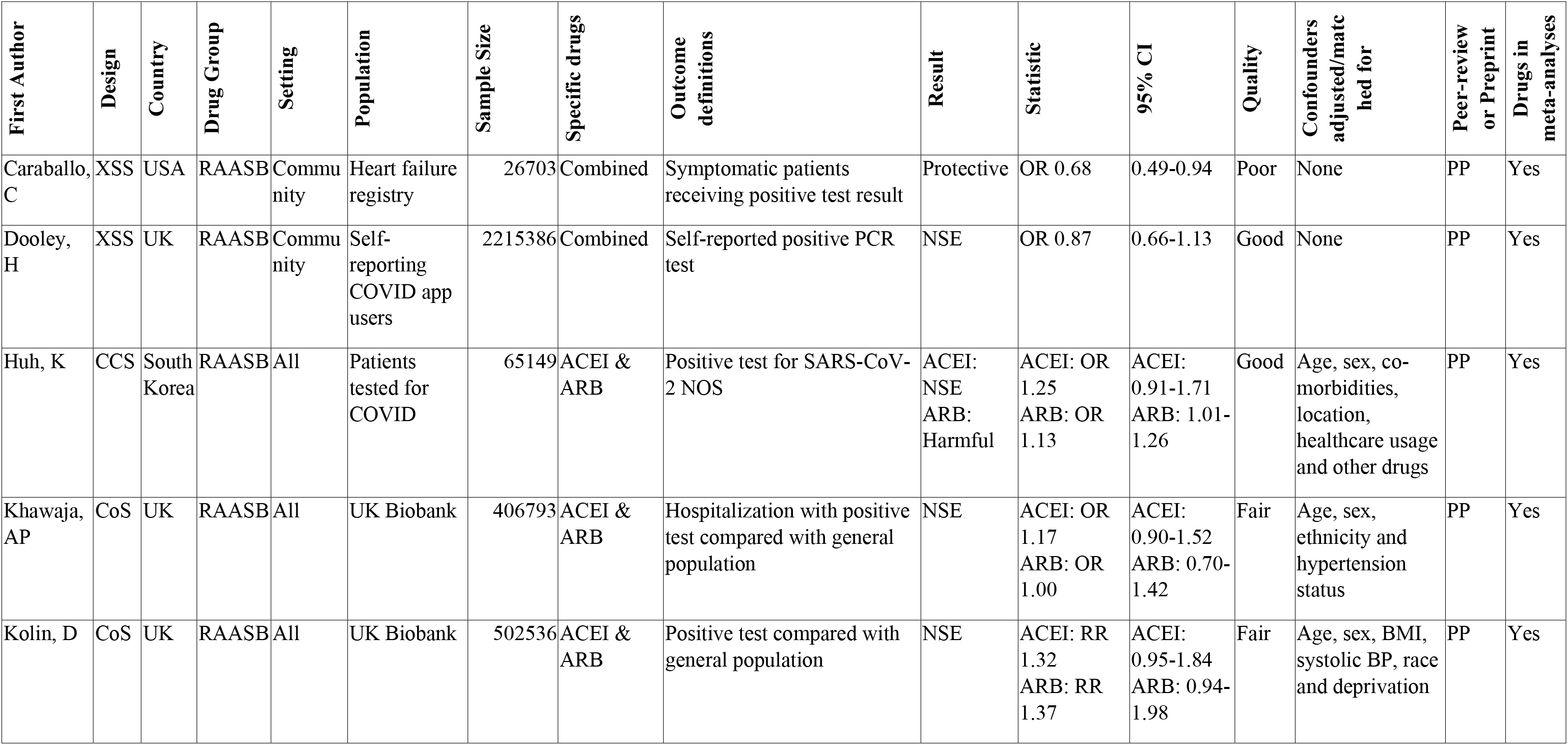
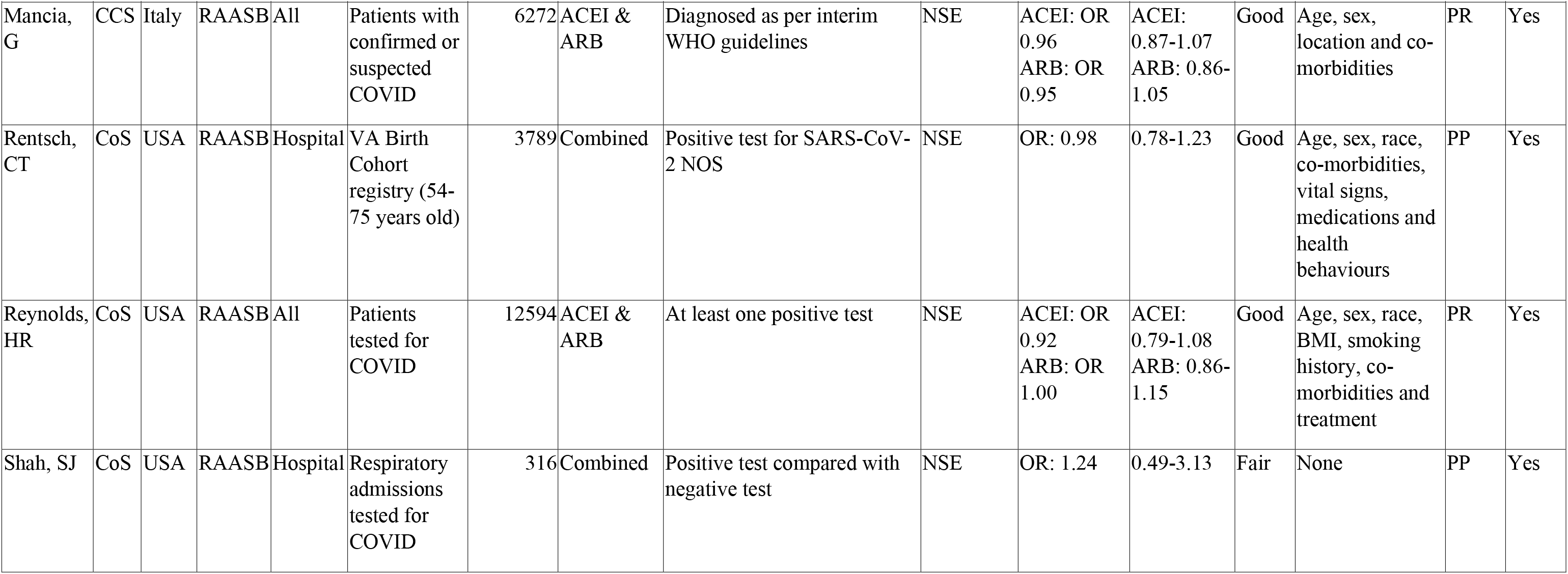
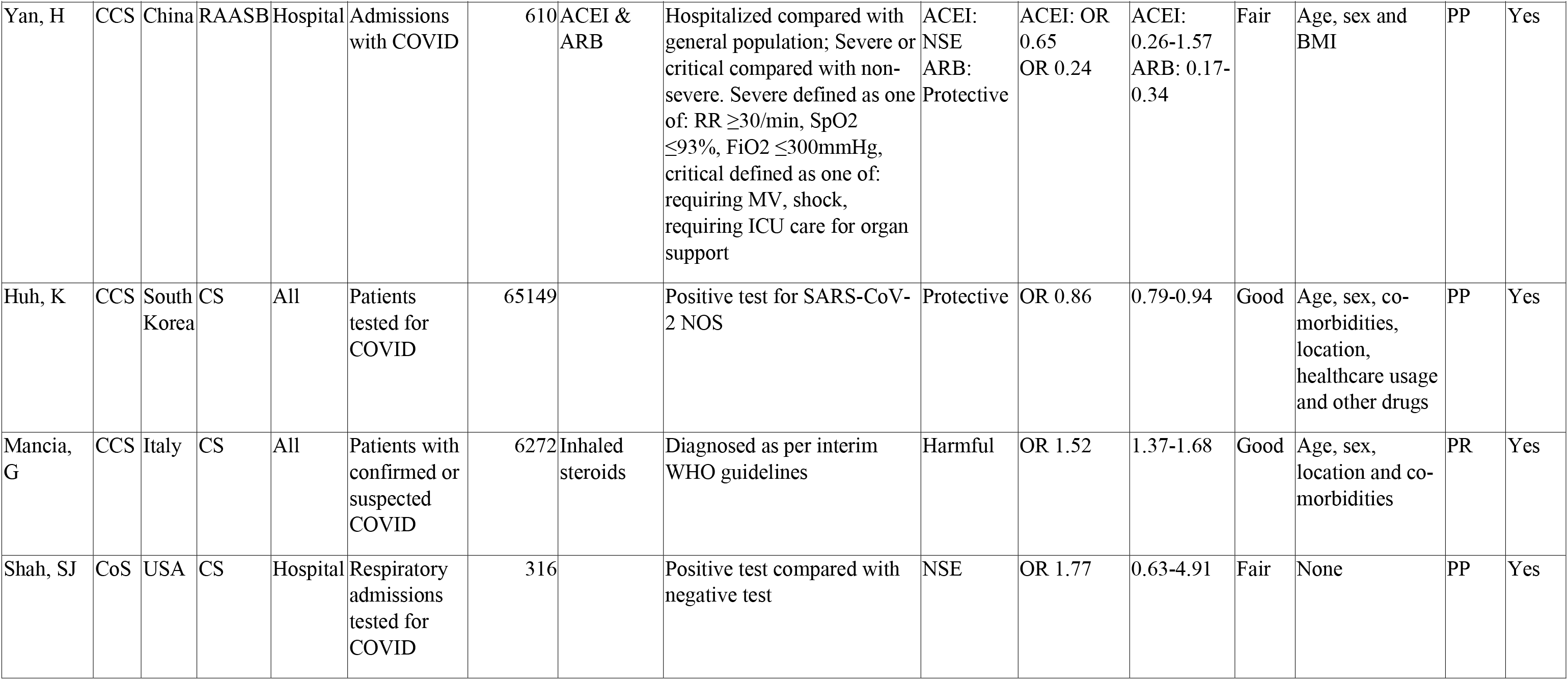
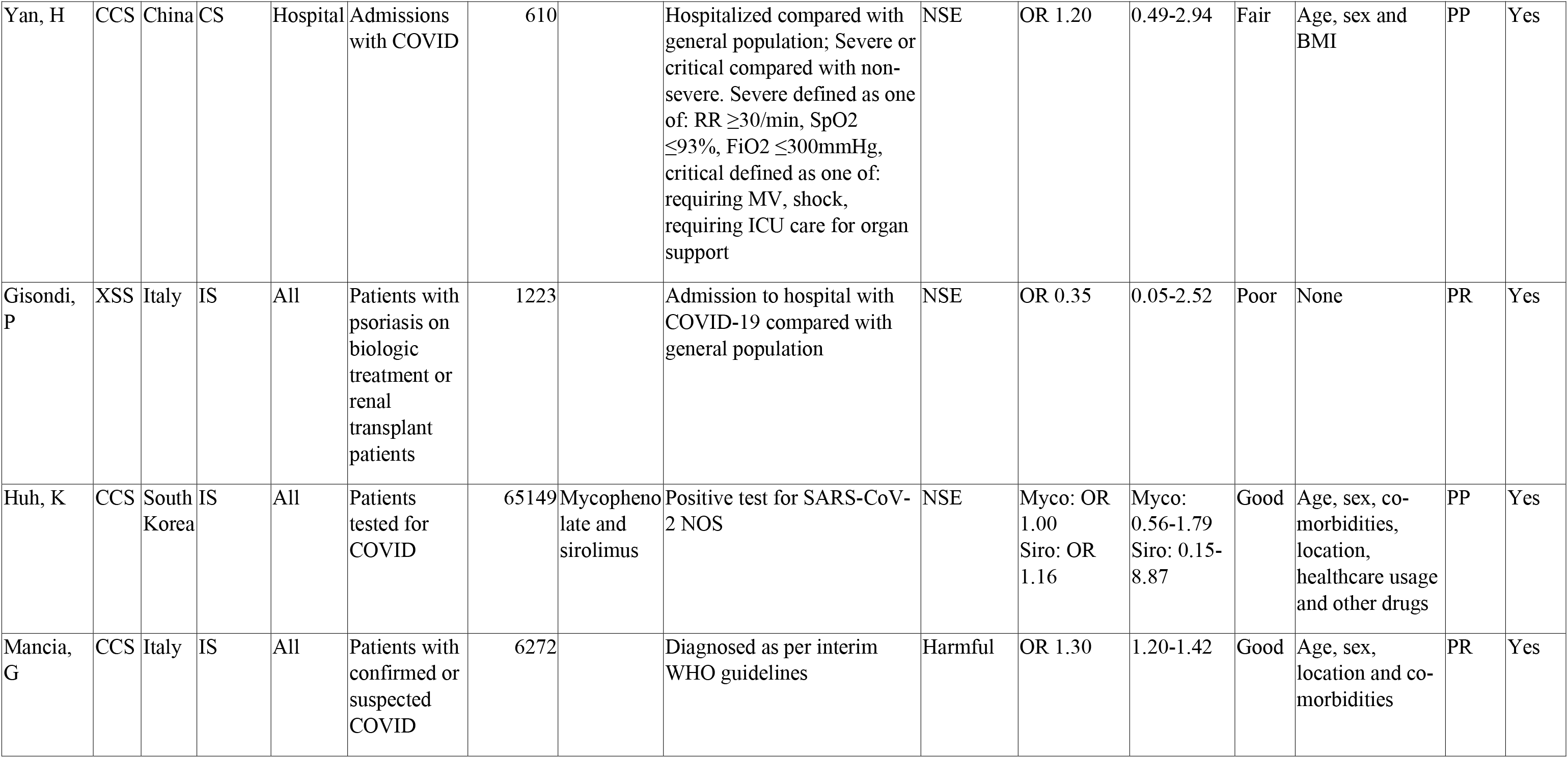
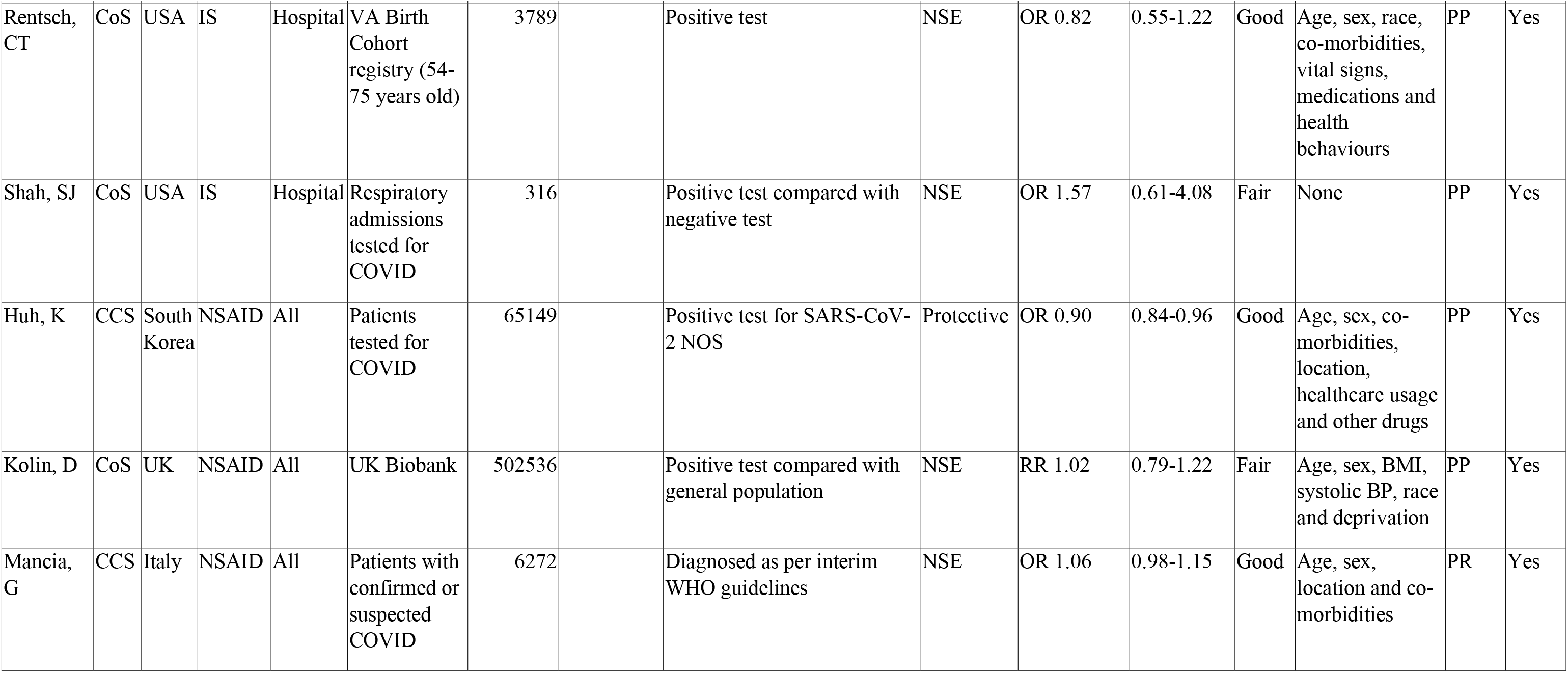
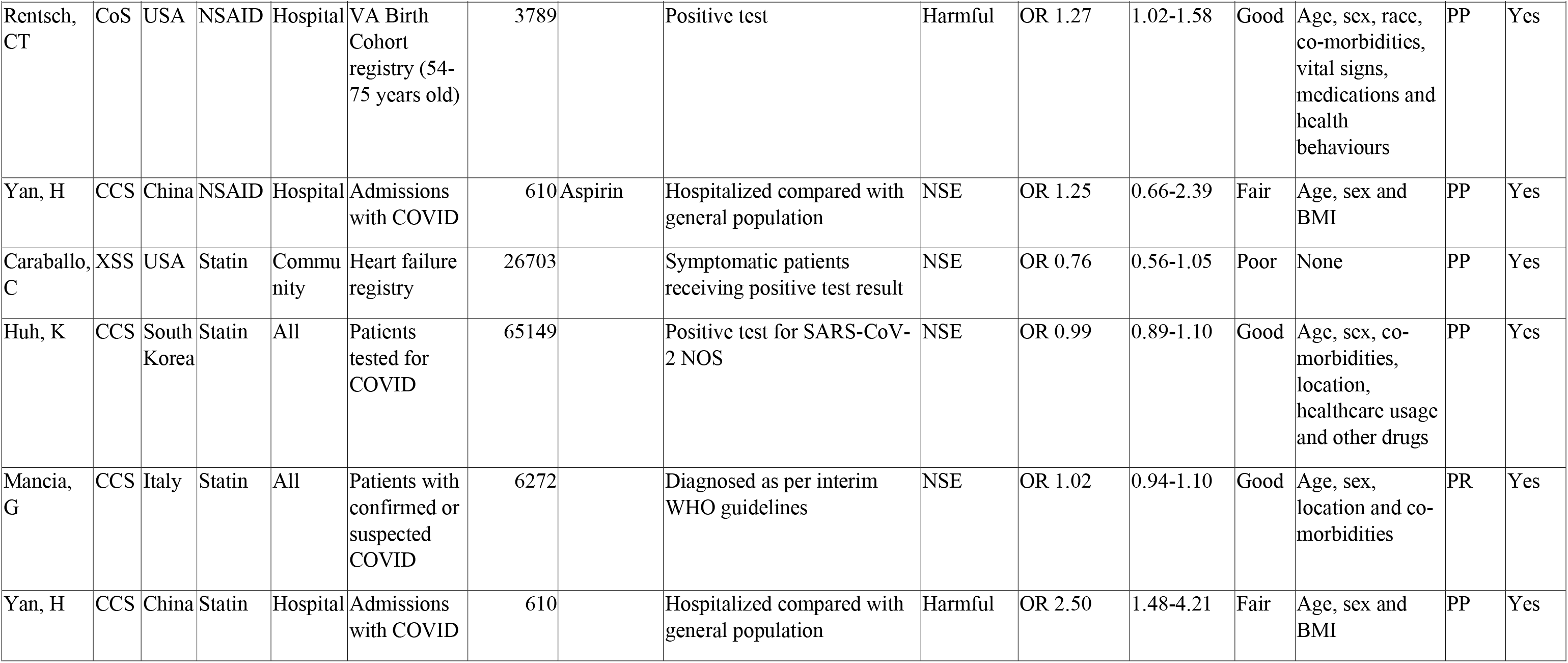
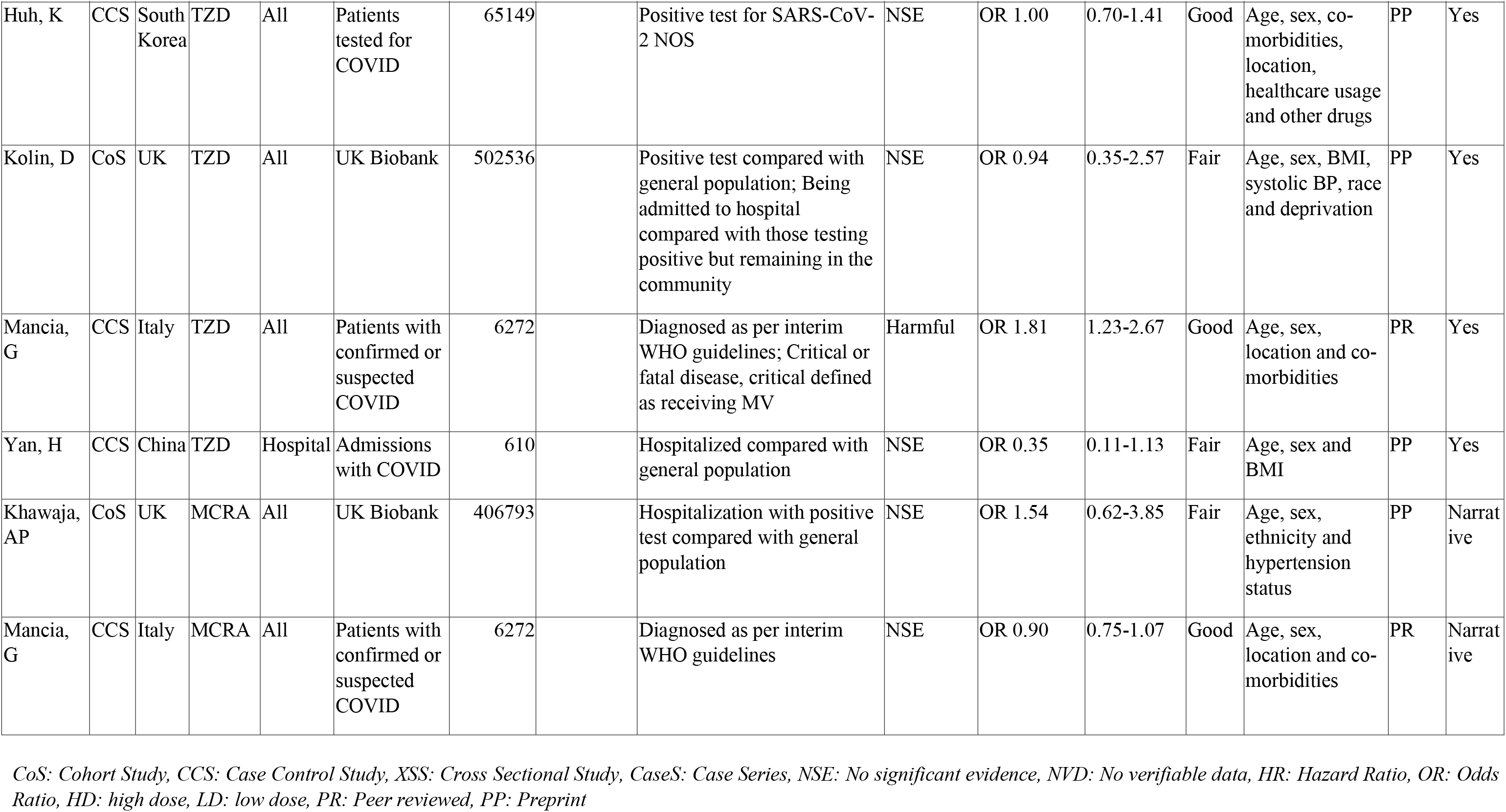
Summary of studies looking into the effect of drug groups on the susceptibility to COVID-19 infection.

| First Author | Design | Country | Drug Group | Setting | Population | Sample Size | Specific drugs | Outcome definitions | Result | Statistic | 95% CI | Quality | Confounders adjusted/matched for | Peer-review or Preprint | Drugs in meta-analyses |
| --- | --- | --- | --- | --- | --- | --- | --- | --- | --- | --- | --- | --- | --- | --- | --- |
| Caraballo, C | XSS | USA | RAASB | Community | Heart failure registry | 26703 | Combined | Symptomatic patients receiving positive test result | Protective | OR 0.68 | 0.49-0.94 | Poor | None | PP | Yes |
| Dooley, H | XSS | UK | RAASB | Community | Self-reporting COVID app users | 2215386 | Combined | Self-reported positive PCR test | NSE | OR 0.87 | 0.66-1.13 | Good | None | PP | Yes |
| Huh, K | CCS | South Korea | RAASB | All | Patients tested for COVID | 65149 | ACEI & ARB | Positive test for SARS-CoV-2 NOS | ACEI: NSE<br>ARB: Harmful | ACEI: OR 1.25<br>ARB: OR 1.13 | ACEI: 0.91-1.71<br>ARB: 1.01-1.26 | Good | Age, sex, co-morbidities, location, healthcare usage and other drugs | PP | Yes |
| Khawaja, AP | CoS | UK | RAASB | All | UK Biobank | 406793 | ACEI & ARB | Hospitalization with positive test compared with general population | NSE | ACEI: OR 1.17<br>ARB: OR 1.00 | ACEI: 0.90-1.52<br>ARB: 0.70-1.42 | Fair | Age, sex, ethnicity and hypertension status | PP | Yes |
| Kolin, D | CoS | UK | RAASB | All | UK Biobank | 502536 | ACEI & ARB | Positive test compared with general population | NSE | ACEI: RR 1.32<br>ARB: RR 1.37 | ACEI: 0.95-1.84<br>ARB: 0.94-1.98 | Fair | Age, sex, BMI, systolic BP, race and deprivation | PP | Yes |

| First Author | Design | Country | Drug Group | Setting | Population | Sample Size | Specific drugs | Outcome definitions | Result | Statistic | 95% CI | Quality | Confounders/adjusted/matched for | Peer-review or Preprint | Drugs in meta-analyses |
| --- | --- | --- | --- | --- | --- | --- | --- | --- | --- | --- | --- | --- | --- | --- | --- |
| Mancia, G | CCS | Italy | RAASB | All | Patients with confirmed or suspected COVID | 6272 | ACEI & ARB | Diagnosed as per interim WHO guidelines | NSE | ACEI: OR 0.96<br>ARB: OR 0.95 | ACEI: 0.87-1.07<br>ARB: 0.86-1.05 | Good | Age, sex, location and co-morbidities | PR | Yes |
| Rentsch, CT | CoS | USA | RAASB | Hospital | VA Birth Cohort registry (54-75 years old) | 3789 | Combined | Positive test for SARS-CoV-2 NOS | NSE | OR: 0.98 | 0.78-1.23 | Good | Age, sex, race, co-morbidities, vital signs, medications and health behaviours | PP | Yes |
| Reynolds, HR | CoS | USA | RAASB | All | Patients tested for COVID | 12594 | ACEI & ARB | At least one positive test | NSE | ACEI: OR 0.92<br>ARB: OR 1.00 | ACEI: 0.79-1.08<br>ARB: 0.86-1.15 | Good | Age, sex, race, BMI, smoking history, co-morbidities and treatment | PR | Yes |
| Shah, SJ | CoS | USA | RAASB | Hospital | Respiratory admissions tested for COVID | 316 | Combined | Positive test compared with negative test | NSE | OR: 1.24 | 0.49-3.13 | Fair | None | PP | Yes |
| Yan, H | CCS | China | RAASB | Hospital | Admissions with COVID | 610 | ACEI & ARB | Hospitalized compared with general population; Severe or critical compared with non-severe. Severe defined as one of: RR ≥30/min, SpO2 ≤93%, FiO2 ≤300mmHg, critical defined as one of: requiring MV, shock, requiring ICU care for organ support | ACEI: NSE<br>ARB: Protective | ACEI: OR 0.65<br>OR 0.24 | ACEI: 0.26-1.57<br>ARB: 0.17-0.34 | Fair | Age, sex and BMI | PP | Yes |
| Huh, K | CCS | South Korea | CS | All | Patients tested for COVID | 65149 |  | Positive test for SARS-CoV-2 NOS | Protective | OR 0.86 | 0.79-0.94 | Good | Age, sex, co-morbidities, location, healthcare usage and other drugs | PP | Yes |
| Mancia, G | CCS | Italy | CS | All | Patients with confirmed or suspected COVID | 6272 | Inhaled steroids | Diagnosed as per interim WHO guidelines | Harmful | OR 1.52 | 1.37-1.68 | Good | Age, sex, location and co-morbidities | PR | Yes |
| Shah, SJ | CoS | USA | CS | Hospital | Respiratory admissions tested for COVID | 316 |  | Positive test compared with negative test | NSE | OR 1.77 | 0.63-4.91 | Fair | None | PP | Yes |
| Yan, H | CCS | China | CS | Hospital | Admissions with COVID | 610 | | Hospitalized compared with general population; Severe or critical compared with non-severe. Severe defined as one of: RR ≥30/min, SpO<sub>2</sub> ≤93%, FiO<sub>2</sub> ≤300mmHg, critical defined as one of: requiring MV, shock, requiring ICU care for organ support | NSE | OR 1.20 | 0.49-2.94 | Fair | Age, sex and BMI | PP | Yes |
| Gisondi, P | XSS | Italy | IS | All | Patients with psoriasis on biologic treatment or renal transplant patients | 1223 |  | Admission to hospital with COVID-19 compared with general population | NSE | OR 0.35 | 0.05-2.52 | Poor | None | PR | Yes |
| Huh, K | CCS | South Korea | IS | All | Patients tested for COVID | 65149 | Mycophenolate and sirolimus | Positive test for SARS-CoV-2 NOS | NSE | Myco: OR 1.00<br>Siro: OR 1.16 | Myco: 0.56-1.79<br>Siro: 0.15-8.87 | Good | Age, sex, co-morbidities, location, healthcare usage and other drugs | PP | Yes |
| Mancia, G | CCS | Italy | IS | All | Patients with confirmed or suspected COVID | 6272 |  | Diagnosed as per interim WHO guidelines | Harmful | OR 1.30 | 1.20-1.42 | Good | Age, sex, location and co-morbidities | PR | Yes |
| Rentsch, CT | CoS | USA | IS | Hospital | VA Birth Cohort registry (54-75 years old) | 3789 |  | Positive test | NSE | OR 0.82 | 0.55-1.22 | Good | Age, sex, race, co-morbidities, vital signs, medications and health behaviours | PP | Yes |
| Shah, SJ | CoS | USA | IS | Hospital | Respiratory admissions tested for COVID | 316 |  | Positive test compared with negative test | NSE | OR 1.57 | 0.61-4.08 | Fair | None | PP | Yes |
| Huh, K | CCS | South Korea | NSAID | All | Patients tested for COVID | 65149 |  | Positive test for SARS-CoV-2 NOS | Protective | OR 0.90 | 0.84-0.96 | Good | Age, sex, co-morbidities, location, healthcare usage and other drugs | PP | Yes |
| Kolin, D | CoS | UK | NSAID | All | UK Biobank | 502536 |  | Positive test compared with general population | NSE | RR 1.02 | 0.79-1.22 | Fair | Age, sex, BMI, systolic BP, race and deprivation | PP | Yes |
| Mancia, G | CCS | Italy | NSAID | All | Patients with confirmed or suspected COVID | 6272 |  | Diagnosed as per interim WHO guidelines | NSE | OR 1.06 | 0.98-1.15 | Good | Age, sex, location and co-morbidities | PR | Yes |
| Rentsch, CT | CoS | USA | NSAID | Hospital | VA Birth Cohort registry (54-75 years old) | 3789 |  | Positive test | Harmful | OR 1.27 | 1.02-1.58 | Good | Age, sex, race, co-morbidities, vital signs, medications and health behaviours | PP | Yes |
| Yan, H | CCS | China | NSAID | Hospital | Admissions with COVID | 610 | Aspirin | Hospitalized compared with general population | NSE | OR 1.25 | 0.66-2.39 | Fair | Age, sex and BMI | PP | Yes |
| Caraballo, C | XSS | USA | Statin | Community | Heart failure registry | 26703 |  | Symptomatic patients receiving positive test result | NSE | OR 0.76 | 0.56-1.05 | Poor | None | PP | Yes |
| Huh, K | CCS | South Korea | Statin | All | Patients tested for COVID | 65149 |  | Positive test for SARS-CoV-2 NOS | NSE | OR 0.99 | 0.89-1.10 | Good | Age, sex, co-morbidities, location, healthcare usage and other drugs | PP | Yes |
| Mancia, G | CCS | Italy | Statin | All | Patients with confirmed or suspected COVID | 6272 |  | Diagnosed as per interim WHO guidelines | NSE | OR 1.02 | 0.94-1.10 | Good | Age, sex, location and co-morbidities | PR | Yes |
| Yan, H | CCS | China | Statin | Hospital | Admissions with COVID | 610 |  | Hospitalized compared with general population | Harmful | OR 2.50 | 1.48-4.21 | Fair | Age, sex and BMI | PP | Yes |
| Huh, K | CCS | South Korea | TZD | All | Patients tested for COVID | 65149 |  | Positive test for SARS-CoV-2 NOS | NSE | OR 1.00 | 0.70-1.41 | Good | Age, sex, co-morbidities, location, healthcare usage and other drugs | PP | Yes |
| Kolin, D | CoS | UK | TZD | All | UK Biobank | 502536 |  | Positive test compared with general population; Being admitted to hospital compared with those testing positive but remaining in the community | NSE | OR 0.94 | 0.35-2.57 | Fair | Age, sex, BMI, systolic BP, race and deprivation | PP | Yes |
| Mancia, G | CCS | Italy | TZD | All | Patients with confirmed or suspected COVID | 6272 |  | Diagnosed as per interim WHO guidelines; Critical or fatal disease, critical defined as receiving MV | Harmful | OR 1.81 | 1.23-2.67 | Good | Age, sex, location and co-morbidities | PR | Yes |
| Yan, H | CCS | China | TZD | Hospital | Admissions with COVID | 610 |  | Hospitalized compared with general population | NSE | OR 0.35 | 0.11-1.13 | Fair | Age, sex and BMI | PP | Yes |
| Khawaja, AP | CoS | UK | MCRA | All | UK Biobank | 406793 |  | Hospitalization with positive test compared with general population | NSE | OR 1.54 | 0.62-3.85 | Fair | Age, sex, ethnicity and hypertension status | PP | Narrative |
| Mancia, G | CCS | Italy | MCRA | All | Patients with confirmed or suspected COVID | 6272 |  | Diagnosed as per interim WHO guidelines | NSE | OR 0.90 | 0.75-1.07 | Good | Age, sex, location and co-morbidities | PR | Narrative |

*CoS: Cohort Study, CCS: Case Control Study, XSS: Cross Sectional Study, CaseS: Case Series, NSE: No significant evidence, NVD: No verifiable data, HR: Hazard Ratio, OR: Odds Ratio, HD: high dose, LD: low dose, PR: Peer reviewed, PP: Preprint*

**Table 4.** Summary of studies looking into the effect of drug groups on the severity of COVID-19 infection.

| First Author | Design | Country | Drug Group | Setting | Population | Sample Size | Acute or Regular | Specific drugs | Outcome definitions | Result | Statistic | 95% CI | Quality | Confounders adjusted/matched for | Peer-review or PP | Drugs in meta-analyses |
| --- | --- | --- | --- | --- | --- | --- | --- | --- | --- | --- | --- | --- | --- | --- | --- | --- |
| Argenziano, MG | CoS | USA | RAASB | Hospital | ED and hospital presentations with COVID | 1000 | Regular | Combined | Transfer to ICU | NSE | OR 1.11 | 0.81-1.53 | Fair | None | PP | Yes |
| Bean, DM | CoS | UK | RAASB | Hospital | Hospital admissions with COVID | 1200 | Regular | Combined | Death or admission to critical care unit for organ support within 21 days of symptom onset | Protective | OR 0.63 | 0.47-0.84 | Good | Co-morbidities, age and sex | PP | Yes |
| Benelli, G | CoS | Italy | RAASB | Hospital | Admissions with COVID | 411 | Regular | ACEIs & ARBs | Requiring NIV or CPAP | ACEI: Harmful<br>ARB: NSE | ACEI: OR 1.95<br>ARB: OR 1.68 | ACEI: 1.06-3.60<br>ARB: 0.94-2.99 | Fair | None | PP | Yes |
| Dauchet, L | XSS | France | RAASB | All | Patients with confirmed or suspected COVID | 288 | Regular | ACEIs & ARBs | ICU admission; Being put through to emergency line due to symptoms | ACEI: NSE<br>ARB: Harmful | ACEI: SPR 0.95<br>ARB: SPR 1.56 | ACEI: 0.55-1.64<br>ARB: 1.02-2.39 | Good | None | PP | Yes |
| Feng, Y | XSS | China | RAASB | Hospital | Admissions with COVID | 476 | Regular | Combined | Critical compared with moderate: critical defined as respiratory failure requiring MV, shock or requiring ICU care. Moderate defined as fever, cough and pneumonia on chest CT but RR<30/min, SpO<sub>2</sub>>93% and FiO<sub>2</sub>>300mmHg (i.e not severe) | Protective | OR 0.17 | 0.05-0.52 | Fair | None | PR | Yes |
| Feng, Z | CoS | China | RAA SB | Hospital | Admissions with COVID | 564 | Regular | Combined | Severe or critical compared with non-severe. Severe defined as one of: RR $\geq 30$/min, SpO2 $\leq 93\%$, FiO2 $\leq 300$mmHg, critical defined as one of: requiring MV, shock, requiring ICU care for organ support | NSE | OR 0.41 | 0.05-3.19 | Good | Propensity score matched and adjusted for age, gender, smoking history, co-morbidities and lung CT score | PP | Yes |
| Kolin, D | CoS | UK | RAA SB | All | UK Biobank | 502536 | Regular | ACEIs & ARBs | Being admitted to hospital compared with those testing positive but remaining in the community | NSE | ACEI: 0.56<br>ARB: 0.60 | ACEI: 0.26-1.22<br>ARB: 0.25-1.42 | Fair | Age, sex, BMI, systolic BP, race and deprivation | PP | Yes |
| Li, J | Case S | China | RAA SB | Hospital | Hypertensive COVID admissions | 1178 | Regular | Combined | Severe compared with non-severe. Severe defined as one of: RR $\geq 30$/min, SpO2 $\leq 93\%$, FiO2 $\leq 300$mmHg, lung infiltrates more than 50% within 24 to 48 hours, respiratory failure and/or multi-organ failure | NSE | OR 1.11 | 0.71-1.73 | Good | None | PR | Yes |
| Li, X | CoS | China | RAA SB | Hospital | Admissions with COVID | 548 | Regular | Combined | Severe or critical compared with non-severe. Severe defined as per IDSA & ATS guidelines for CAP | NSE | OR 0.86 | 0.45-1.61 | Good | Age, sex, blood tests, antiviral usage | PR | Yes |
| Liu, Yingxia | CoS | China | RAA SB | Hospital | Hypertensive COVID admissions | 78 | Regular | ACEIs & ARBs | Severe compared with non-severe as per NHCPRC 2019-nCoV guidelines | NSE | ACEI: OR 0.57<br>ARB: OR 0.54 | ACEI: 0.11-2.95<br>ARB: 0.25-1.16 | Fair | Sex | PP | Yes |
| Mancia, G | CCS | Italy | RAA SB | All | Patients with confirmed or suspected COVID | 6272 | Regular | ACEIs & ARBs | Critical or fatal disease, critical defined as receiving MV | NSE | ACEI: OR 0.91<br>ARB: OR 0.83 | ACEI: 0.69-1.21<br>ARB: 0.63-1.10 | Good | Age, sex, location and co-morbidities | PR | Yes |
| Meng, J | CoS | China | RAA<br>SB | Hospital | Hypertensive COVID admissions | 42 | Regular | Combined | Severe disease or death, severe defined as per NHCPRC 2019-nCoV guidelines | NSE | OR 0.28 | 0.07-1.12 | Fair | None | PR | Yes |
| Rentsch, CT | CoS | USA | RAA<br>SB | Hospital | VA Birth Cohort registry (54-75 years old) | 3789 | Regular | Combined | Hospitalization compared with community care after testing positive | NSE | OR 1.15 | 0.71-1.87 | Good | Age, sex, race, co-morbidities, vital signs, medications and health behaviours | PP | Yes |
| Reynolds, HR | CoS | USA | RAA<br>SB | All | Patients tested for COVID | 12594 | Regular | ACEIs & ARBs | ICU admission, MV or death | NSE | ACEI: OR 0.90<br>ARB: OR 0.99 | ACEI: 0.70-1.14<br>ARB: 0.79-1.25 | Good | Age, sex, race, BMI, smoking history, co-morbidities and treatment | PR | Yes |
| Yan, H | CCS | China | RAA<br>SB | Hospital | Admissions with COVID | 610 | Regular | ACEIs & ARBs | Severe or critical compared with non-severe. Severe defined as one of: RR $\geq 30/\text{min}$, SpO2 $\leq 93\%$, FiO2 $\leq 300\text{mmHg}$, critical defined as one of: requiring MV, shock, requiring ICU care for organ support | NSE | ACEI: OR 1.23<br>ARB: OR 0.77 | ACEI: 0.19-7.93<br>ARB: 0.36-1.63 | Fair | Age, sex and BMI | PP | Yes |
| Yang, G | CoS | China | RAA<br>SB | Hospital | Hypertensive COVID admissions | 126 | Regular | Combined | Severe or critical compared with non-severe. Severe defined as one of: RR $\geq 30/\text{min}$, SpO2 $\leq 93\%$, FiO2 $\leq 300\text{mmHg}$, critical defined as one of: requiring MV, shock, requiring ICU care for organ support | NSE | OR 0.73 | 0.34-1.58 | Fair | None | PP | Yes |
| Zeng, Z | CoS | China | RAA<br>SB | Hospital | Hypertensive COVID Admissions | 75 | Regular | Combined | Severe compared with non-severe. Severe defined as per IDSA & ATS guidelines for CAP | NSE | OR 2.46 | 0.94-6.45 | Poor | None | PP | Yes |
| Argenziano, MG | CoS | USA | CS | Hospital | ED and hospital presentations with COVID | 1000 | Regular | Inhaled, nasal, oral steroids | Transfer to ICU | NSE | OR 1.06 | 0.59-1.90 | Fair | None | PP | Yes |
| Cao, C | CoS | China | CS | Hospital | Admissions with mild-moderate COVID | 58 | Acute | Methylprednisolone | Severe compared with non-severe. Severe defined as one of: RR ≥ 33/min, SpO<sub>2</sub> ≤ 94% at rest, PO<sub>2</sub>/FiO<sub>2</sub> ≤ 300mmHg, requiring MV, developing shock or multiple organ failure requiring ICU | Protective | OR 0.14 | 0.02-0.80 | Fair | None | PP | Narrative |
| Li, X | CoS | China | CS | Hospital | Admissions with COVID | 548 | Regular | Oral | Severe or critical compared with non-severe. Severe defined as per IDSA & ATS guidelines for CAP | NSE | Oral: OR 0.51<br>Inh: OR 0.69 | Oral: 0.09-2.84<br>Inh: 0.11-4.16 | Good | Age, sex, blood tests, antiviral usage | PR | Yes |
| Liu, Yanli | CoS | China | CS | Hospital | Admissions with COVID | 109 | Acute |  | Berlin definition of ARDS | NSE | OR 1.38 | 0.64-2.98 | Fair | None | PP | Narrative |
| Lui, GCY | CoS | China | CS | Hospital | Admissions with COVID | 114 | Acute | Methylprednisolone | ICU admission, MV or death | Harmful | HR 4.17 | 1.23-14.11 | Poor | Age, sex, co-morbidities and blood markers | PP | OR incalculable or inaccurate |
| Robilotti, EV | CoS | USA | CS | Hospital | Cancer patients with symptomatic COVID | 423 | Regular | Steroids or lymphopaenia | Requiring high-flow oxygen or MV | NSE | OR 1.52 | 0.92-2.50 | Fair | Age, race, treatment and co-morbidities | PP | Yes |
| Wang, D | CoS | China | CS | Hospital | Admissions with COVID | 115 | Acute | Methylprednisolone | ICU admission or death | NSE | OR 2.16 | 0.49-9.43 | Poor | Age, sex, co-morbidities and blood markers | PP | Narrative |
| Yan, H | CCS | China | CS | Hospital | Admissions with COVID | 610 | Regular | | Hospitalized compared with general population; Severe or critical compared with non-severe. Severe defined as one of: RR $\geq 30$/min, SpO<sub>2</sub> $\leq 93\%$, FiO<sub>2</sub> $\leq 300$mmHg, critical defined as one of: requiring MV, shock, requiring ICU care for organ support | Harmful | OR 7.56 | 1.17-48.93 | Fair | Age, sex and BMI | PP | Yes |
| Yi, P | CoS | China | CS | Hospital | Admissions with COVID | 100 | Acute | | Severe or critical compared with non-severe. Severe defined as one of: RR $\geq 30$/min, SpO<sub>2</sub> $\leq 93\%$, FiO<sub>2</sub> $\leq 300$mmHg, critical defined as one of: requiring MV, shock, requiring ICU care for organ support | NSE | - | - | Fair | Age and BMI | PP | OR incalculable or inaccurate |
| Li, X | CoS | China | IS | Hospital | Admissions with COVID | 548 | Regular |  | Severe or critical compared with non-severe. Severe defined as per IDSA & ATS guidelines for CAP | NSE | OR 0.35 | 0.11-4.16 | Good | Age, sex, blood tests, antiviral usage | PR | Yes |
| Monreal, E | CoS | Spain | IS | Hospital | Admissions with COVID | 138 | Regular |  | Berlin definition for ARDS | Protective | OR 0.16 | 0.05-0.52 | Fair | Age, sex and time of onset | PP | Yes |
| Rentsch, CT | CoS | USA | IS | Hospital | VA Birth Cohort registry (54-75 years old) | 3789 | Regular |  | Hospitalization compared with community care after testing positive | NSE | OR 1.62 | 0.75-3.50 | Good | Age, sex, race, comorbidities, vital signs, medications and health behaviours | PP | Yes |
| Robilotti, EV | CoS | USA | IS | Hospital | Cancer patients with symptomatic COVID | 423 | Regular | Chemotherapy | Requiring high-flow oxygen or MV | NSE | OR 1.16 | 0.76-1.78 | Fair | Age, race, treatment and co-morbidities | PP | Yes |
| Roumier, M | CCS | France | IS | Hospital | Admissions with COVID | 60 | Acute | Tocilizumab | Requiring MV | Protective | OR 0.42 | 0.20-0.89 | Fair | Matched for age, sex and severity | PP | No |
| Zhu, L | CCS | China | IS | Hospital | Renal transplant admissions with COVID | 10 | Regular |  | Severe compared with non-severe. Severe defined as per 7th edition of NHCPRC 7th version; Time from onset to shedding (unclear); Recovery compared with death | Harmful | - | - | Fair | None | PR | OR incalculable or inaccurate |
| Argenziano, MG | CoS | USA | NSAID | Hospital | ED and hospital presentations with COVID | 1000 | Regular |  | Transfer to ICU | NSE | OR 0.78 | 0.55-1.11 | Fair | None | PP | Yes |
| Castro, VM | XSS | USA | NSAID | Community | Patients tested for COVID | 2271 | Regular | Ibuprofen, naproxen, ketorolac | Being admitted to hospital compared with those testing positive but remaining in the community | Protective and harmful | Ibu: OR 0.65<br>Nap: OR 0.39<br>Ket: OR 1.70 | Ibu: 0.42-0.99<br>Nap: 0.14-0.93<br>Ket: 1.03-2.78 | Good | Age, sex, race, Charlston index, location | PP | Yes |
| Hong, W | CoS | China | NSAID | Hospital | Admissions with COVID | 44 | Acute | Celecoxib | Progression to severe or critical disease. Severe defined as one of: RR $\geq 30$/min, SpO2 $\leq 93\%$, FiO2 $\leq 300$mmHg. Critical defined as one of: requiring MV, shock, requiring ICU care for organ support | NSE | - | - | Poor | None | PP | No |
| Kolin, D | CoS | UK | NSAID | All | UK Biobank | 502536 | Regular |  | Being admitted to hospital compared with those testing positive but remaining in the community | NSE | RR 0.75 | 0.47-1.18 | Fair | Age, sex, BMI, systolic BP, race and deprivation | PP | Yes |
| Kragholm, K | CCS | Denmark | NSAID | All | National registry (>30 years old without heart failure) | 1872 | Regular | Ibuprofen | Severe disease, ICU admission or death. Severe disease defined as ICD-10 code for severe adult respiratory syndrome | NSE | OR 1.57 | 0.72-3.38 | Fair | Age, sex and co-morbidities | PP | Yes |
| Rentsch, CT | CoS | USA | NSAID | Hospital | VA Birth Cohort registry (54-75 years old) | 3789 | Regular |  | Hospitalization compared with community care after testing positive | Harmful | OR 1.18 | 0.74-1.89 | Good | Age, sex, race, co-morbidities, vital signs, medications and health behaviours | PP | Yes |
| Yan, H | CCS | China | NSAID | Hospital | Admissions with COVID | 610 | Regular | Aspirin | Severe or critical compared with non-severe. Severe defined as one of: RR $\geq 30/\text{min}$, $\text{SpO}_2 \leq 93\%$, $\text{FiO}_2 \leq 300\text{mmHg}$, critical defined as one of: requiring MV, shock, requiring ICU care for organ support | Protective and harmful | OR 0.51 | 0.12-2.13 | Fair | Age, sex and BMI | PP | Yes |
| Argenziano, MG | CoS | USA | Statin | Hospital | ED and hospital presentations with COVID | 1000 | Regular |  | Transfer to ICU | NSE | OR 1.20 | 0.89-1.63 | Fair | None | PP | Narrative |
| Yan, H | CCS | China | Statin | Hospital | Admissions with COVID | 610 | Regular | | Severe or critical compared with non-severe. Severe defined as one of: RR $\geq 30/\text{min}$, $\text{SpO}_2 \leq 93\%$, $\text{FiO}_2 \leq 300\text{mmHg}$, critical defined as one of: requiring MV, shock, requiring ICU care for organ support | Harmful | OR 0.98 | 0.32-2.99 | Fair | Age, sex and BMI | PP | Narrative |
| Yan, H | CCS | China | TZD | Hospital | Admissions with COVID | 610 | Regular | | Severe or critical compared with non-severe. Severe defined as one of: RR $\geq 30/\text{min}$, SpO2 $\leq 93\%$, FiO2 $\leq 300\text{mmHg}$, critical defined as one of: requiring MV, shock, requiring ICU care for organ support | NSE | OR 4.52 | 0.34-59.54 | Fair | Age, sex and BMI | PP | Yes |

CoS: Cohort Study, CCS: Case Control Study, XSS: Cross Sectional Study, CaseS: Case Series, NSE: No significant evidence, NVD: No verifiable data, HR: Hazard Ratio, OR: Odds Ratio, HD: high dose, LD: low dose, PR: Peer reviewed, PP: Preprint

**Table 5.** Summary of studies looking into the effect of drug groups on the mortality from COVID-19 infection.

| **First Author** | **Design** | **Country** | **Drug Group** | **Setting** | **Population** | **Sample Size** | **Acute or Regular** | **Outcome definitions** | **Result** | **Statistic** | **95% CI** | **Quality** | **Confounders adjusted/ matched for** | **Peer-review or Preprint** | **Drugs in meta-analyses** |
| --- | --- | --- | --- | --- | --- | --- | --- | --- | --- | --- | --- | --- | --- | --- | --- |
| Benelli, G | CoS | Italy | RAA SB | Hospital | Admissions with COVID | 411 | Regular | Deaths compared with discharges and inpatients | NSE | ACEI: OR 1.39<br>ARB: OR 1.54 | ACEI: 0.67-2.86<br>ARB: 0.79-2.98 | Fair | None | PP | Yes |
| Chen, Ming | CCS | China | RAA SB | Hospital | Deaths and discharges from hospital with COVID | 123 | Regular | Deaths compared with discharges | NSE | OR 1.13 | 0.28-4.54 | Fair | None | PP | Yes |
| Guo, T | XSS | China | RAA SB | Hospital | Admissions with COVID | 187 | Regular | Deaths compared with discharges | NSE | OR 1.70 | 0.63-4.58 | Fair | None | PR | Yes |
| Ip, A | CCS | USA | RAA SB | Hospital | Hypertensive COVID discharges and deaths | 1216 | Regular | Deaths compared with discharges | Protective | OR 0.66 | 0.51-0.85 | Poor | None | PP | Yes |
| Lee, HY | CoS | South Korea | RAA SB | All | Patients with lab confirmed COVID | 8266 | Regular | Mortality within 60 days | NSE | HR 1.07 | 0.68-1.65 | Good | Age, sex and co-morbidities | PP | Narrative |
| Li, J | CaseS | China | RAA SB | Hospital | Hypertensive COVID admissions | 1178 | Regular | In hospital death compared with discharges | NSE | OR 0.76 | 0.44-1.33 | Good | None | PR | Yes |
| Richardson, S | CaseS | USA | RAA SB | Hospital | Hypertensive COVID discharges and deaths | 5700 | Regular | In hospital death compared with discharges | NSE | ACEI: OR 1.34<br>ARB: OR 1.21 | ACEI: 0.94-1.91<br>ARB: 0.89-1.65 | Fair | None | PR | Yes |
| Sánchez - Álvarez, JE | XSS | Spain | RAA SB | All | Renal replacement patients with COVID | 375 | Regular | In hospital death compared with cured cases | Protective | OR 0.59 | 0.39-0.90 | Poor | None | PR | Yes |

| First Author | Design | Country | Drug Group | Setting | Population | Sample Size | Acute or Regular | Outcome definitions | Result | Statistic | 95% CI | Quality | Confounders adjusted/ matched for | Peer-review or Preprint | Drugs in meta-analyses |
| --- | --- | --- | --- | --- | --- | --- | --- | --- | --- | --- | --- | --- | --- | --- | --- |
| Yang, G | CoS | China | RAA SB | Hospital | Hypertensive COVID admissions | 126 | Regular | Deaths compared with discharges and inpatients | NSE | OR 0.32 | 0.07-1.51 | Fair | None | PP | Yes |
| Zhang, P | CoS | China | RAA SB | Hospital | Hypertensive COVID admissions | 1128 | Regular | 28 day mortality | Protective | HR 0.37 | 0.15-0.89 | Good | Matched for age, sex, symptoms, co-morbidities, CT findings, CRP and creatinine. Adjusted for location, treatment, D-Dimer and unilateral lesion | PR | Narrative |
| Cao, J | CoS | China | CS | Hospital | Admissions with COVID | 102 | Acute | All deaths occurring in data collection period compared with those discharged | NSE | OR 2.06 | 0.70-6.09 | Fair | None | PR | Yes |
| Chen, Ming | CCS | China | CS | Hospital | Deaths and discharges from hospital with COVID | 123 | Acute | Deaths compared with discharges | Harmful | OR 4.75 | 1.78-12.66 | Fair | None | PP | Yes |
| Li, X | CoS | China | CS | Hospital | Admissions with COVID | 548 | Acute | Deaths at any point from admission until final date of follow up (4 weeks after last admission) | HD: Harmful<br>LD: NSE | HD: HR 3.5<br>LD: HR 1.26 | HD: 1.79-6.86<br>LD: 0.61-2.58 | Good | Age, sex, blood tests, antiviral usage | PR | Narrative |
| Liang, M | CoS/ CCS | China | CS | Hospital | Admissions with COVID | 120 | Acute | Death during hospital stay | NSE | OR 0.65 | 0.10-4.14 | Good | None | PP | Yes |
| Liu, Yanli | CoS | China | CS | Hospital | Admissions with COVID | 109 | Acute | Deaths compared with discharges | NSE | - | - | Fair | None | PP | OR incalculable or inaccurate |
| Lu, X | CoS | China | CS | ICU | Critical COVID discharges and deaths | 244 | Acute | 28 day mortality | NSE | - | - | Good | Propensity score matched and adjusted for age, SpO2 and lymphocyte count | PP | OR incalculable or inaccurate |
| Sánchez - Álvarez, JE | XSS | Spain | CS | All | Renal replacement patients with COVID | 375 | Acute | In hospital death compared with cured cases | Harmful | OR 1.81 | 1.12-2.91 | Poor | None | PR | Yes |
| Wu, C | CoS | China | CS | Hospital | COVID patients with ARDS | 201 | Acute | Death at any point up to 18 days after recruitment | Protective | HR 0.38 | 0.20-0.72 | Fair | None | PR | Yes |
| Zhang, S | CoS | China | CS | Hospital | Admissions with COVID | 262 | Acute | 28 day mortality | Harmful | OR 4.97 | 2.85-8.45 | Poor | None | PP | Yes |
| Zhou, F | CoS | China | CS | Hospital | Admissions with COVID | 191 | Acute | Deaths compared with discharges | Harmful | OR 3.18 | 1.63-6.19 | Poor | None | PR | Yes |
| Monreal, E | CoS | Spain | IS | Hospital | Admissions with COVID | 138 | Regular | Deaths compared with those with a final outcome | NSE |  |  | Fair | Age, sex and time of onset | PP | Yes |
| Roumier, M | CCS | France | IS | Hospital | Admissions with COVID | 60 | Acute | In-hospital deaths within 2 days of end of recruitment period | NSE |  |  | Fair | Matched for age, sex and severity | PP | No |

CoS: Cohort Study, CCS: Case Control Study, XSS: Cross Sectional Study, CaseS: Case Series, NSE: No significant evidence, NVD: No verifiable data, HR: Hazard Ratio, OR: Odds Ratio, HD: high dose, LD: low dose, PR: Peer reviewed, PP: Preprint

## Appendix 4: funnel plots

**Fig 12.**
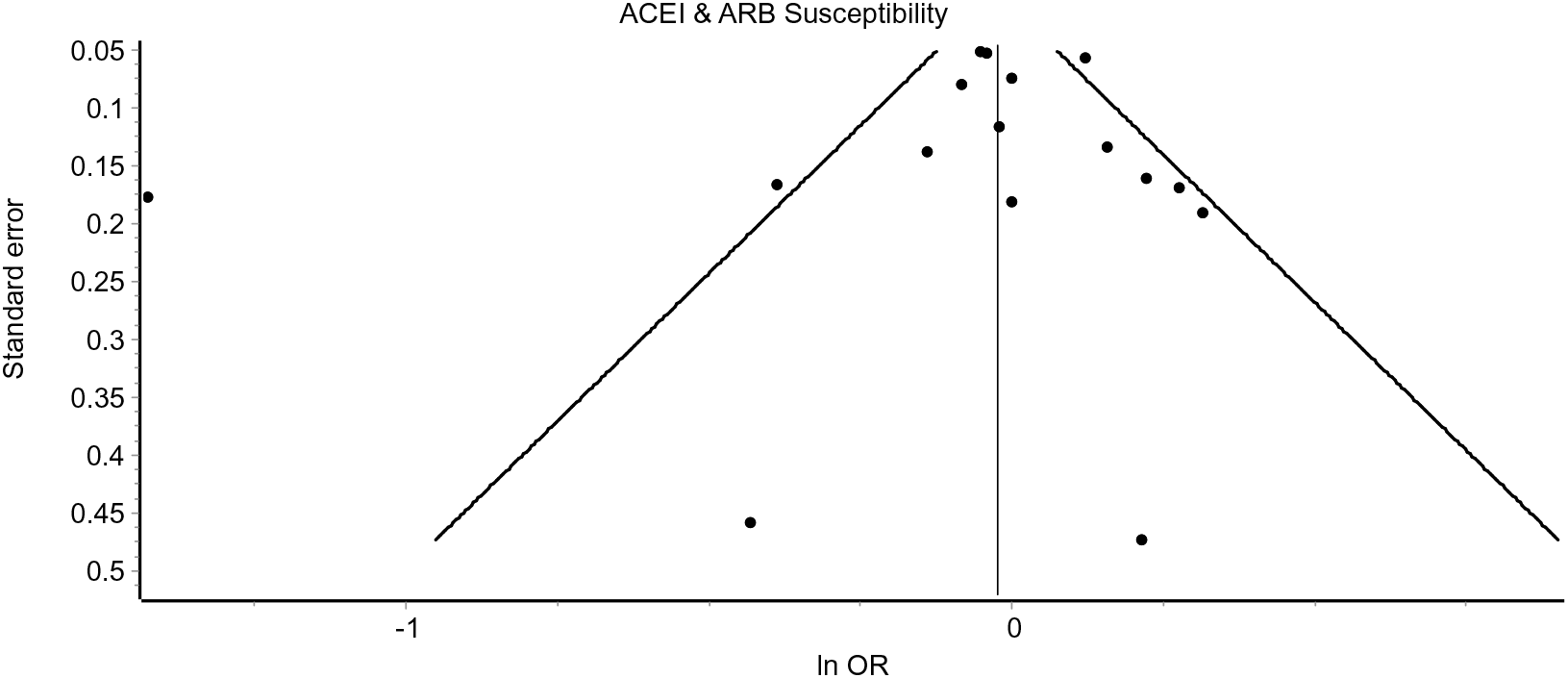
Funnel plot to assess for publication bias from studies measuring susceptibility to COVID-19 in those taking RAASBs. ACEI – angiotensin-converting enzyme inhibitor, ARB – angiotensin-II-receptor blocker, OR – odds ratio.

**Fig 13.**
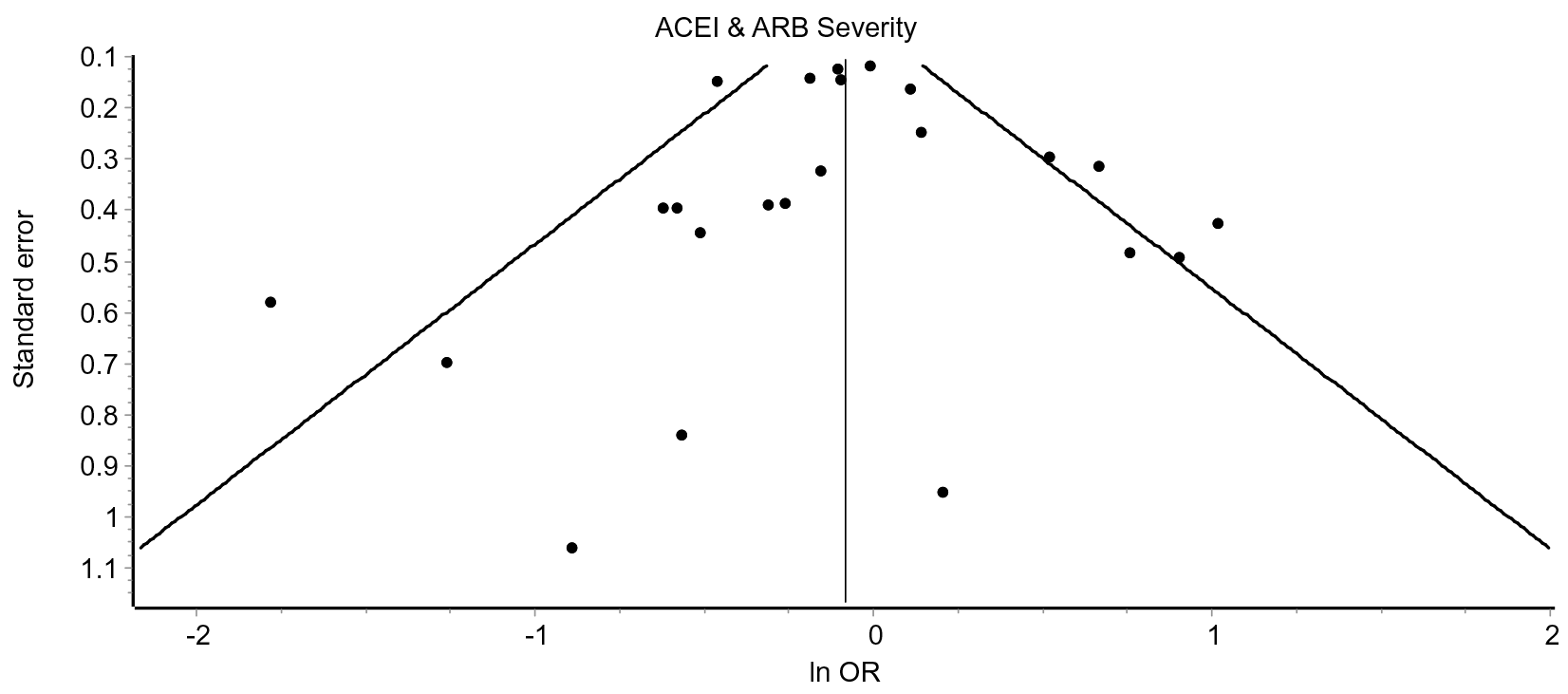
Funnel plot to assess for publication bias from studies measuring severity of COVID-19 in those taking RAASBs. ACEI – angiotensin-converting enzyme inhibitor, ARB – angiotensin-II-receptor blocker, OR – odds ratio.

## Appendix 5: data collection table headings

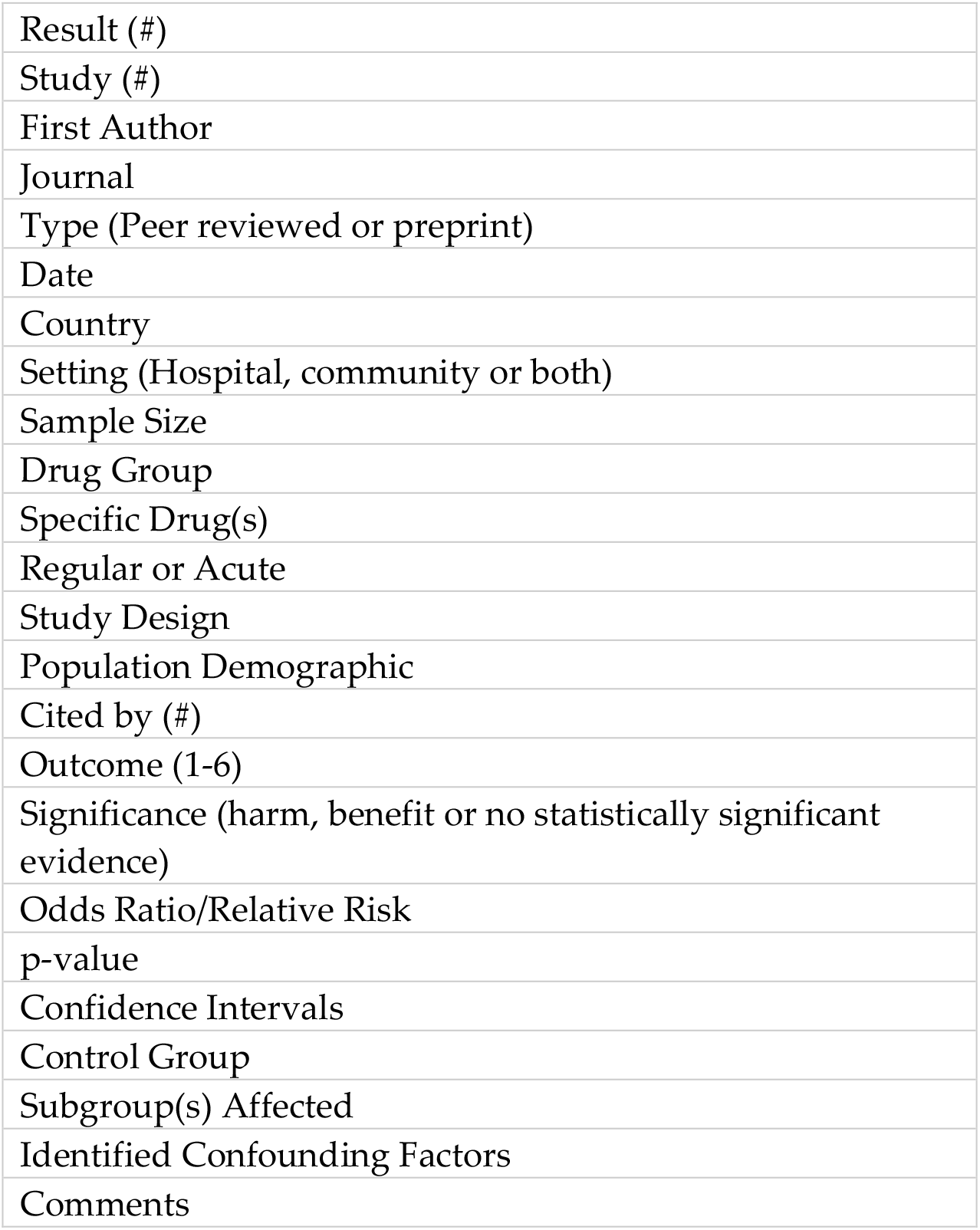

## Appendix 6: forest plots

**Fig 14.**
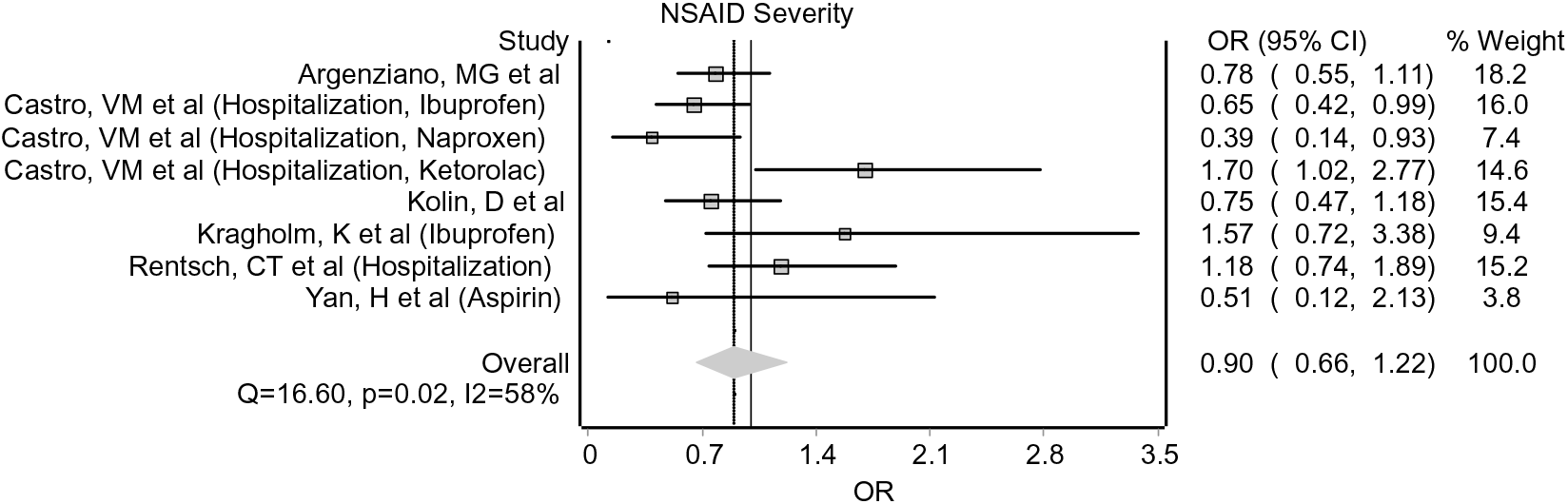
Forest plot for effect of NSAIDs on severity of COVID-19. NSAID – non-steroidal anti-inflammatory drug, OR – odds ratio, CI – confidence interval.

**Fig 15.**
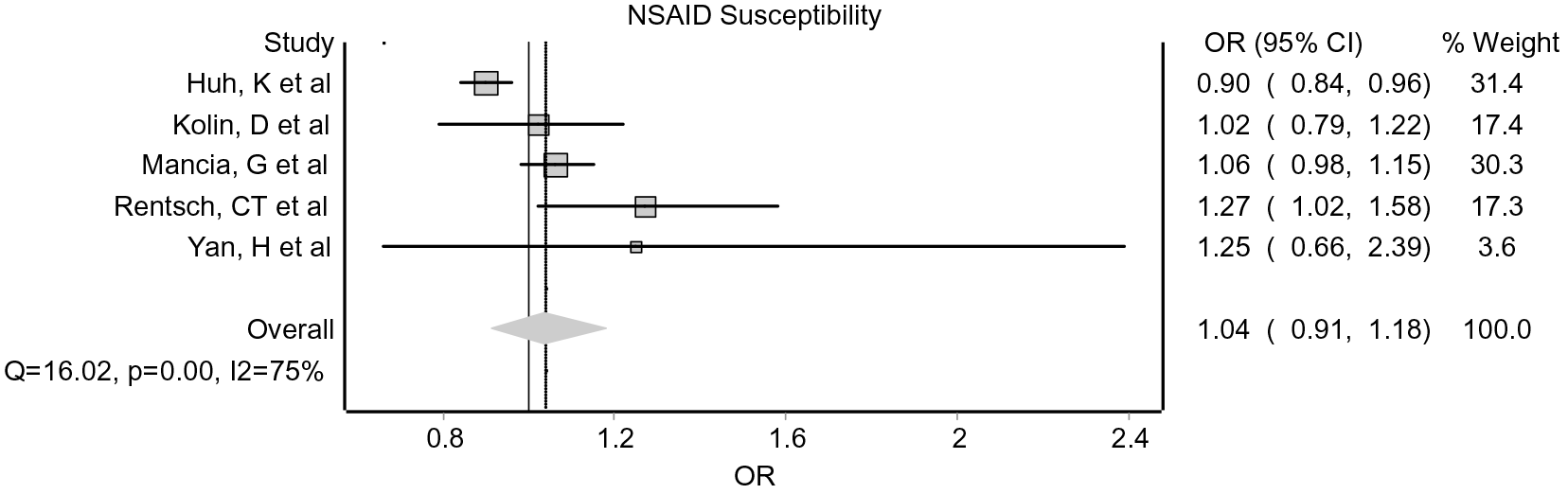
Forest plot for effect of NSAIDs on susceptibility to COVID-19. NSAID – non-steroidal anti-inflammatory drug, OR – odds ratio, CI – confidence interval.

**Fig 16.**
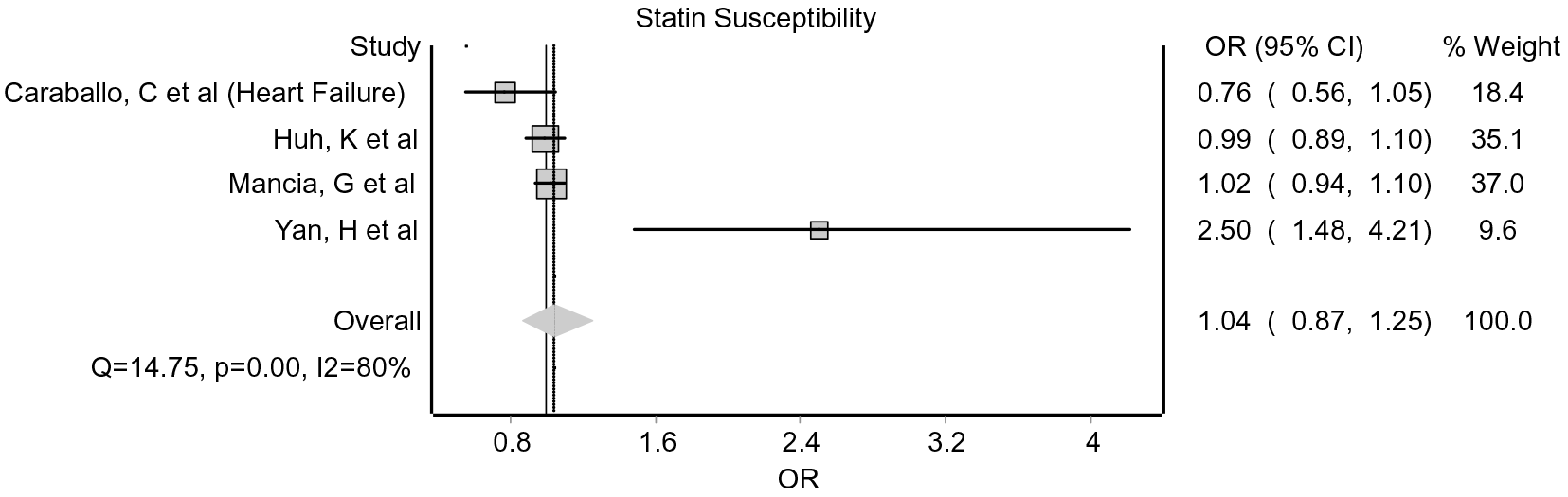
Forest plot for effect of statins on susceptibility to COVID-19. OR – odds ratio, CI – confidence interval.

**Fig 17.**
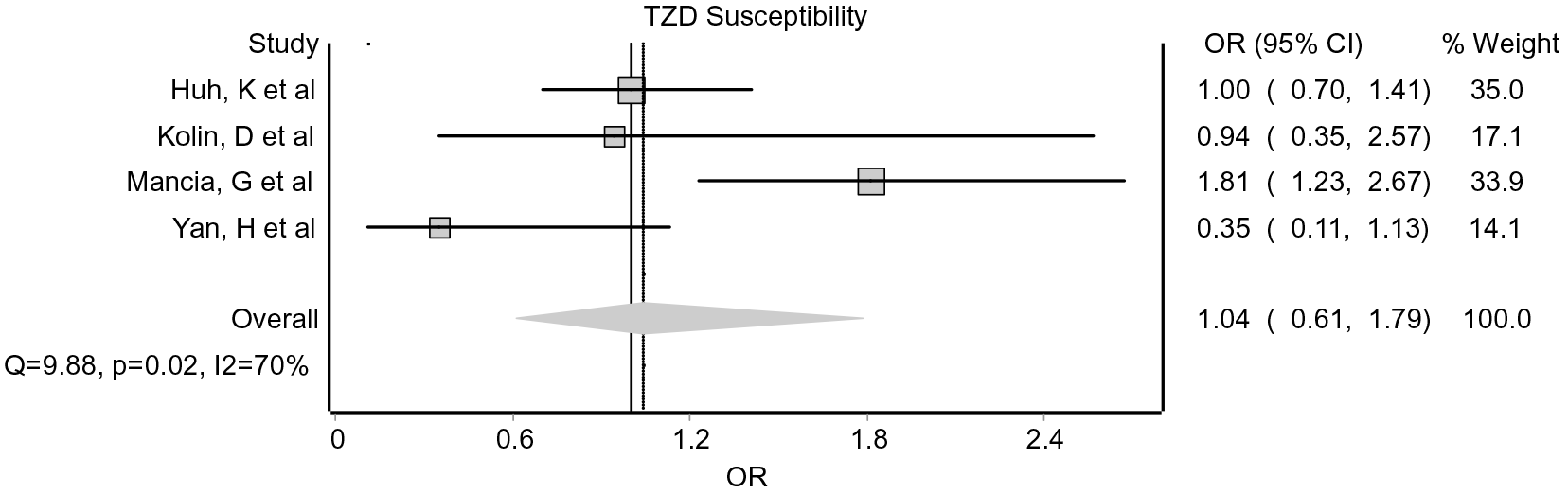
Forest plot for effect of TZDs on susceptibility to COVID-19. TZD - thiazolidinediones, OR – odds ratio, CI – confidence interval.

**Fig 18.**
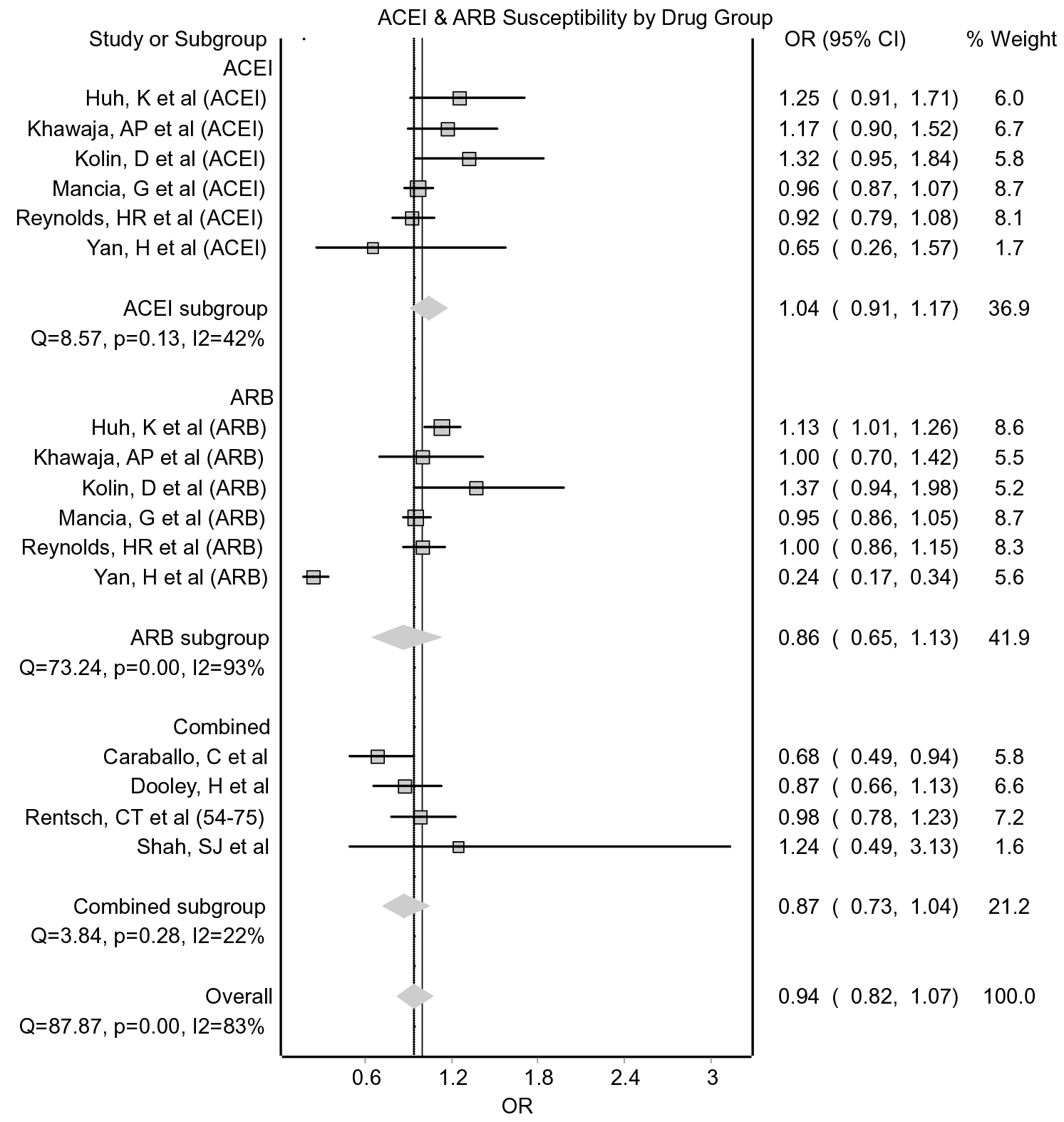
Forest plot of susceptibility to COVID-19 for those taking regular RAAS blockers, split by drug group. ACEI – angiotensin-converting enzyme inhibitor, ARB – angiotensin-II-receptor blocker, OR – odds ratio, CI – confidence interval.

**Fig 19.**
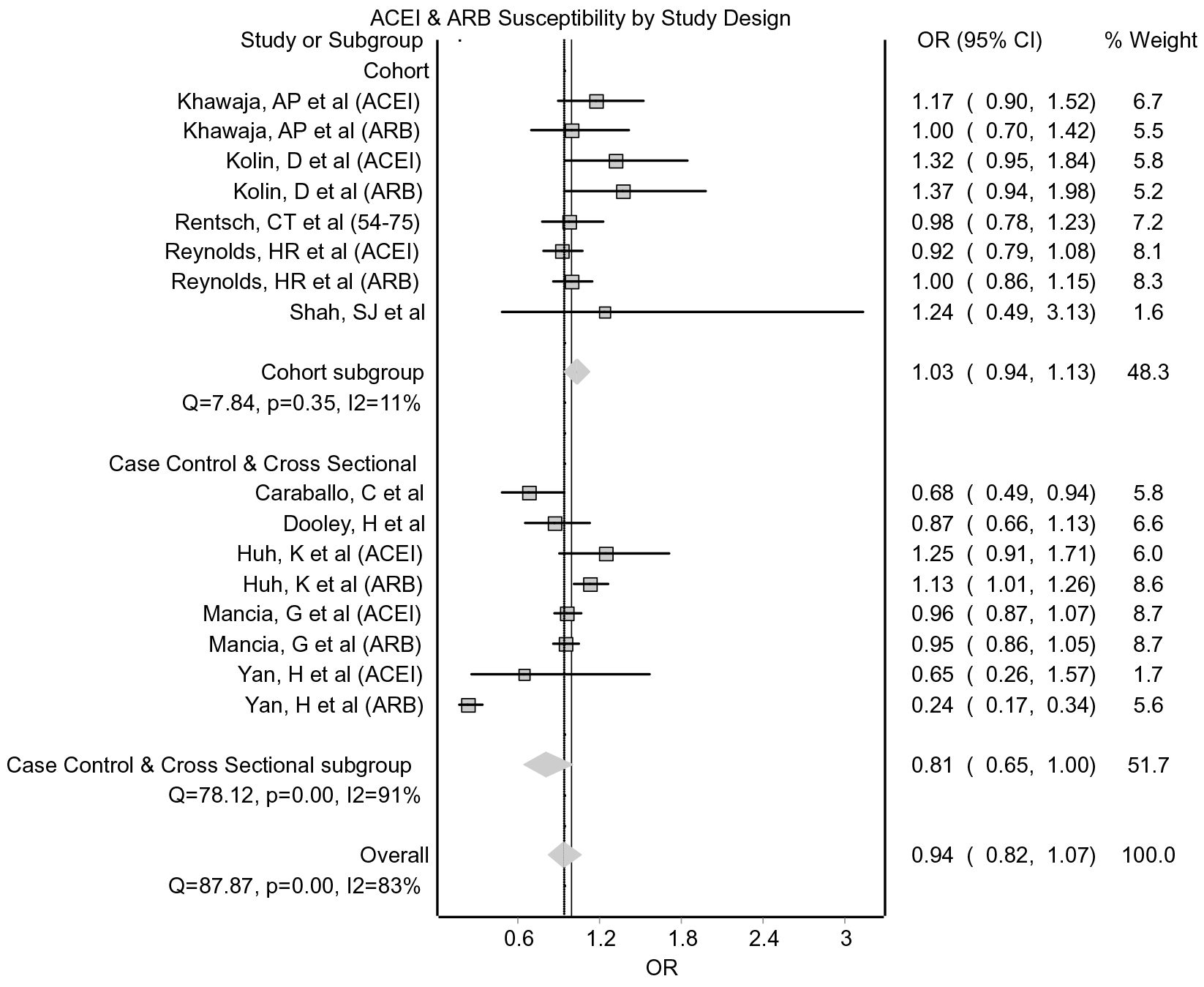
Forest plot of susceptibility to COVID-19 for those taking regular RAAS blockers, split by study design. ACEI – angiotensin-converting enzyme inhibitor, ARB – angiotensin-II-receptor blocker, OR – odds ratio, CI – confidence interval.

**Fig 20.**
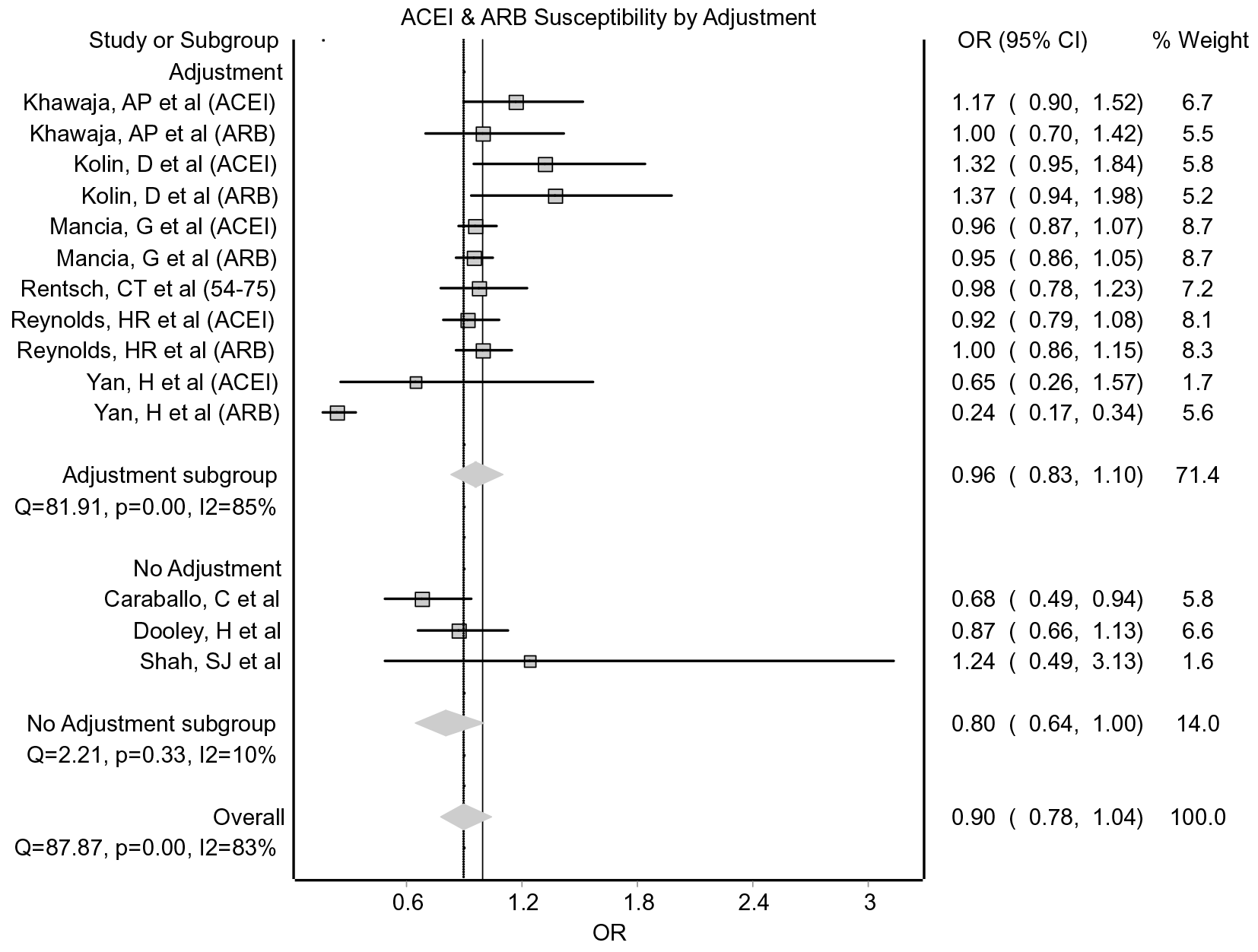
Forest plot of susceptibility to COVID-19 for those taking regular RAAS blockers, split by level of adjustment. ACEI – angiotensin-converting enzyme inhibitor, ARB – angiotensin-II-receptor blocker, OR – odds ratio, CI – confidence interval.

## Appendix 7: sensitivity analysis for removal of preprints

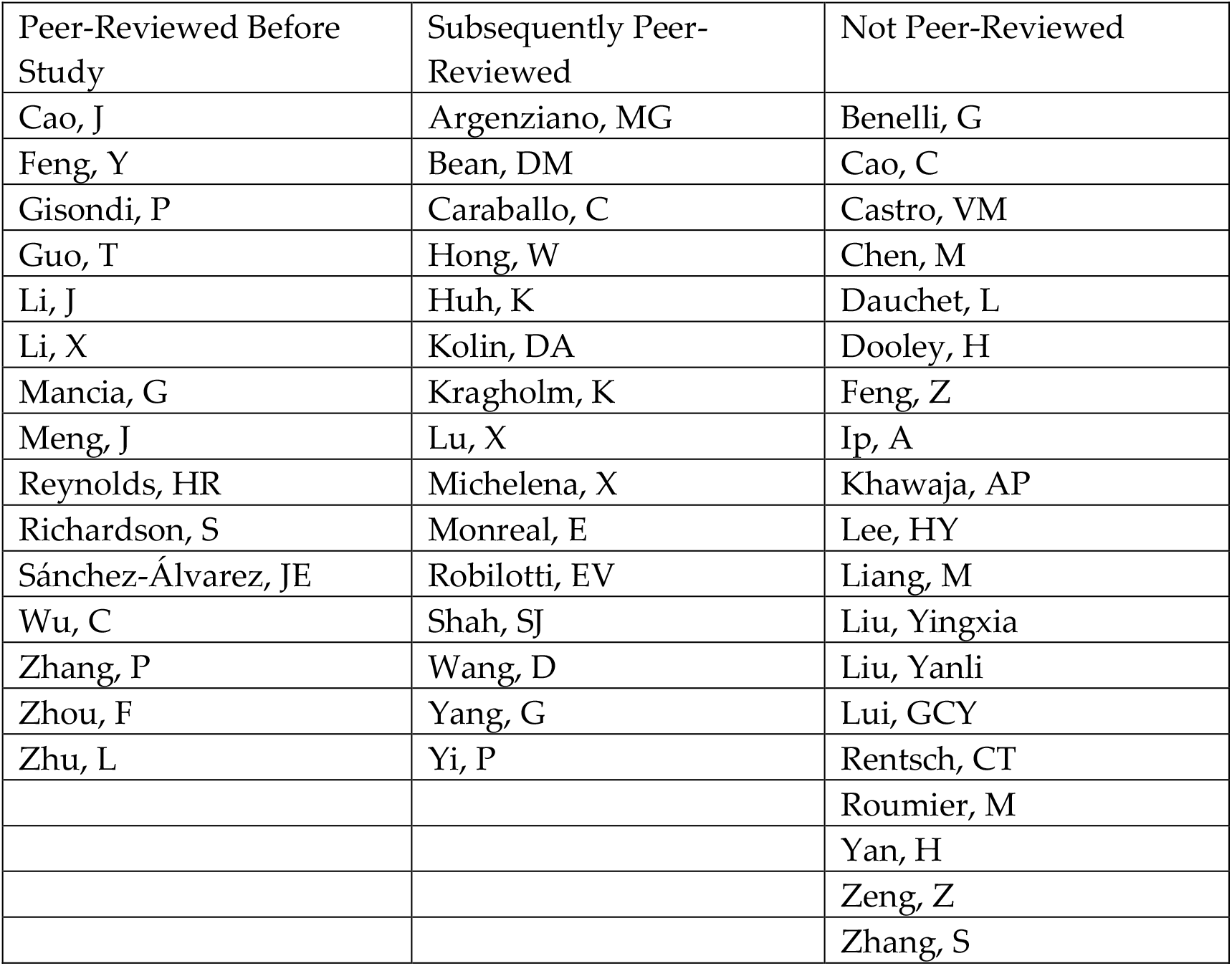

### RAAS blockers

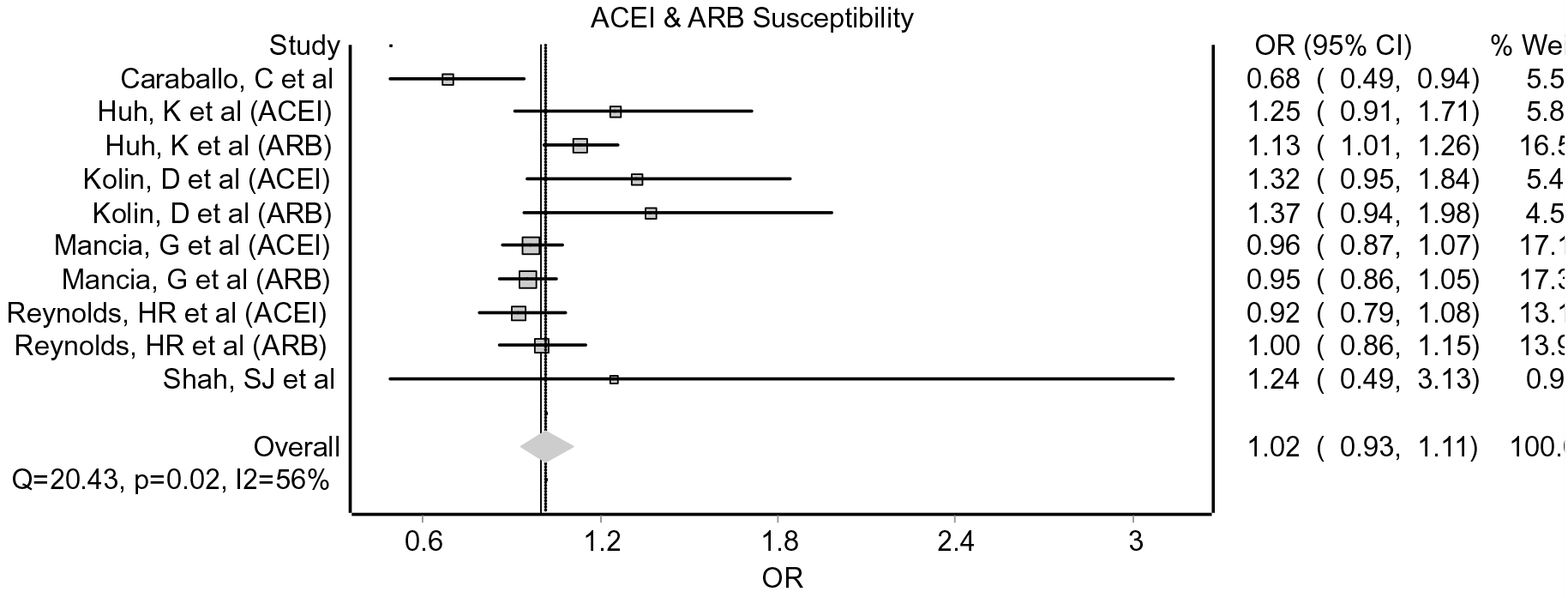
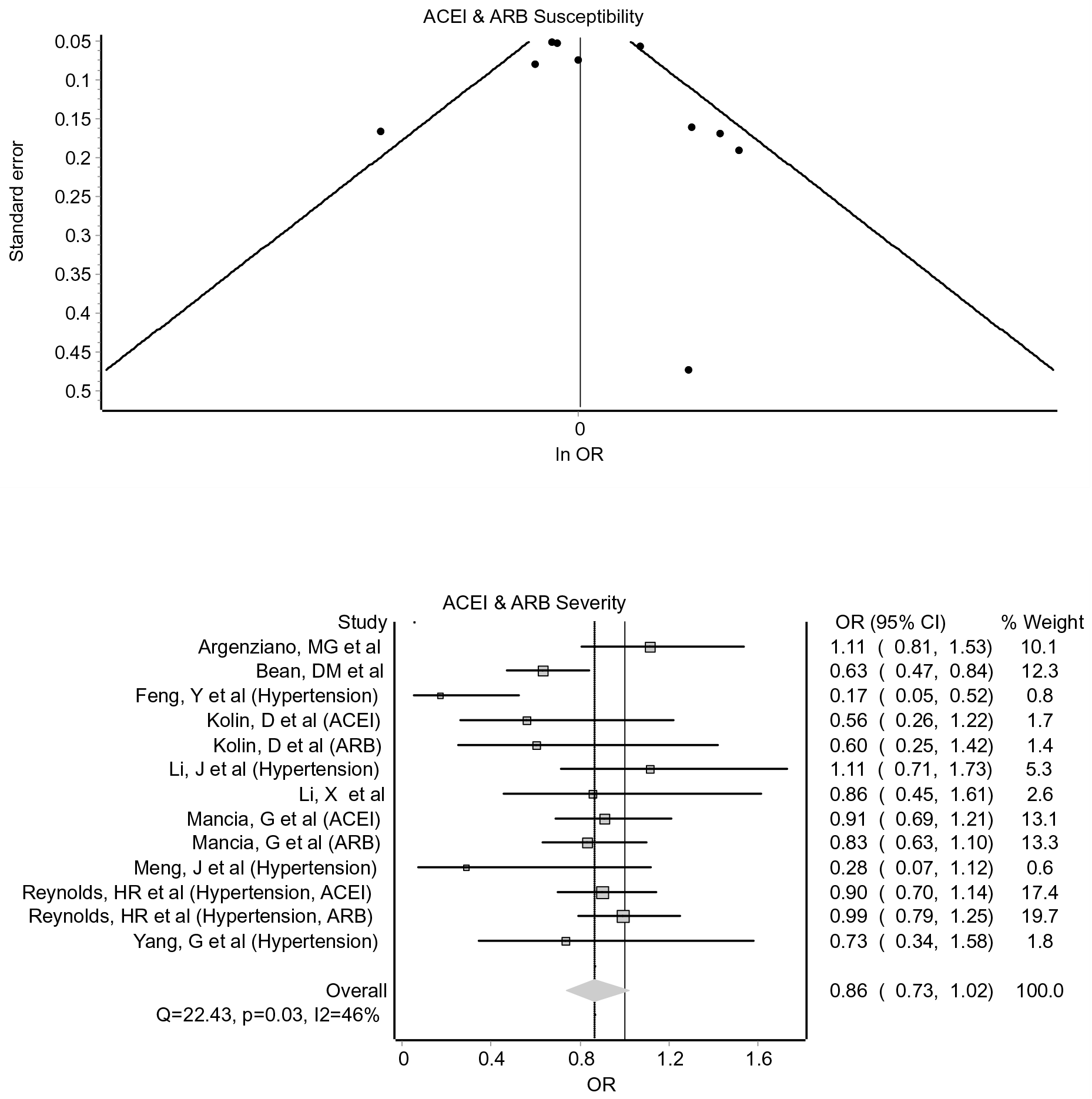
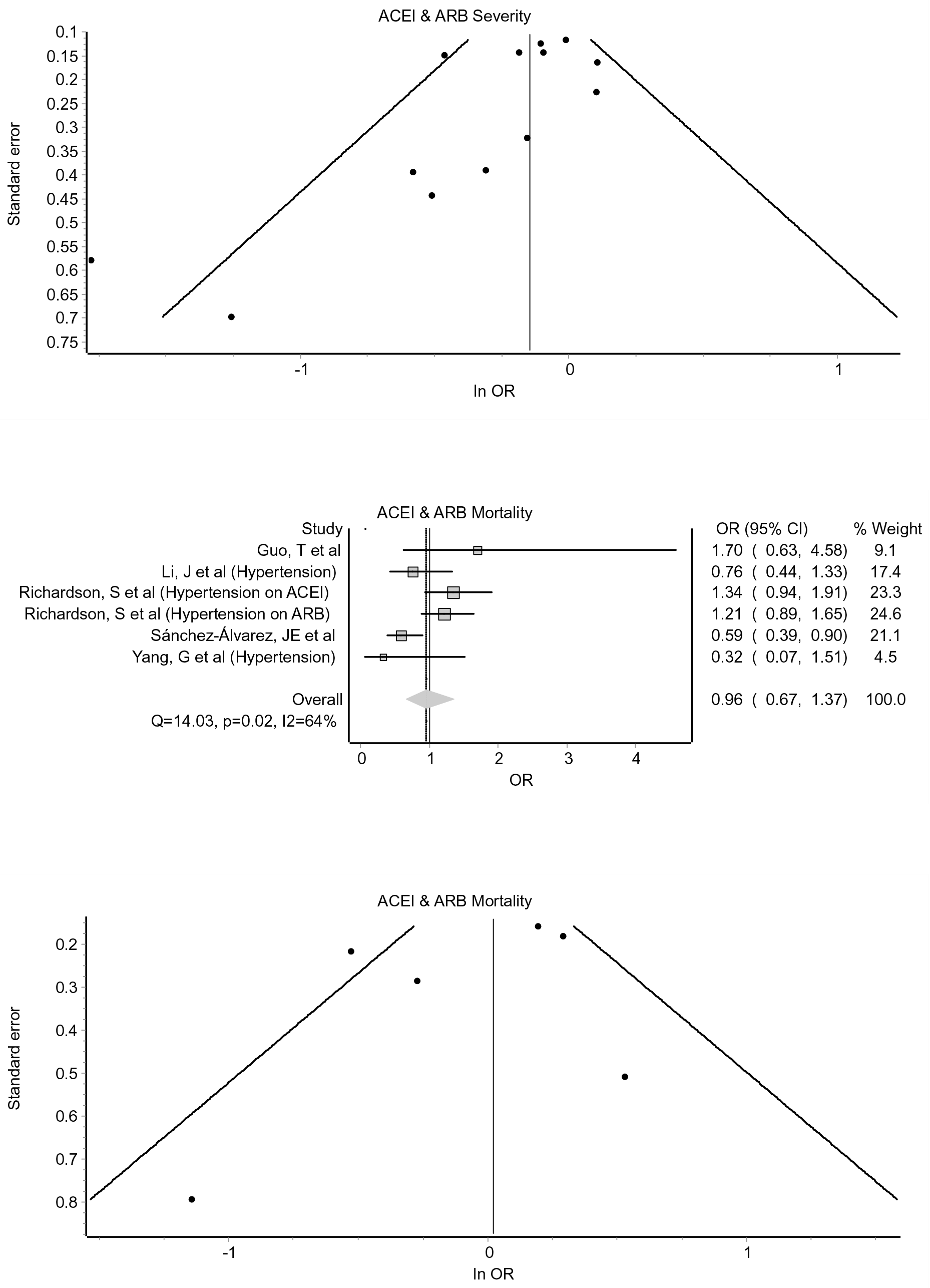

### Corticosteroids

No meta-analysis was performed for susceptibility with corticosteroids. Severity with acute corticosteroids leaves Wang, D only: OR 2.16 (95% CI 0.49 –9.43).

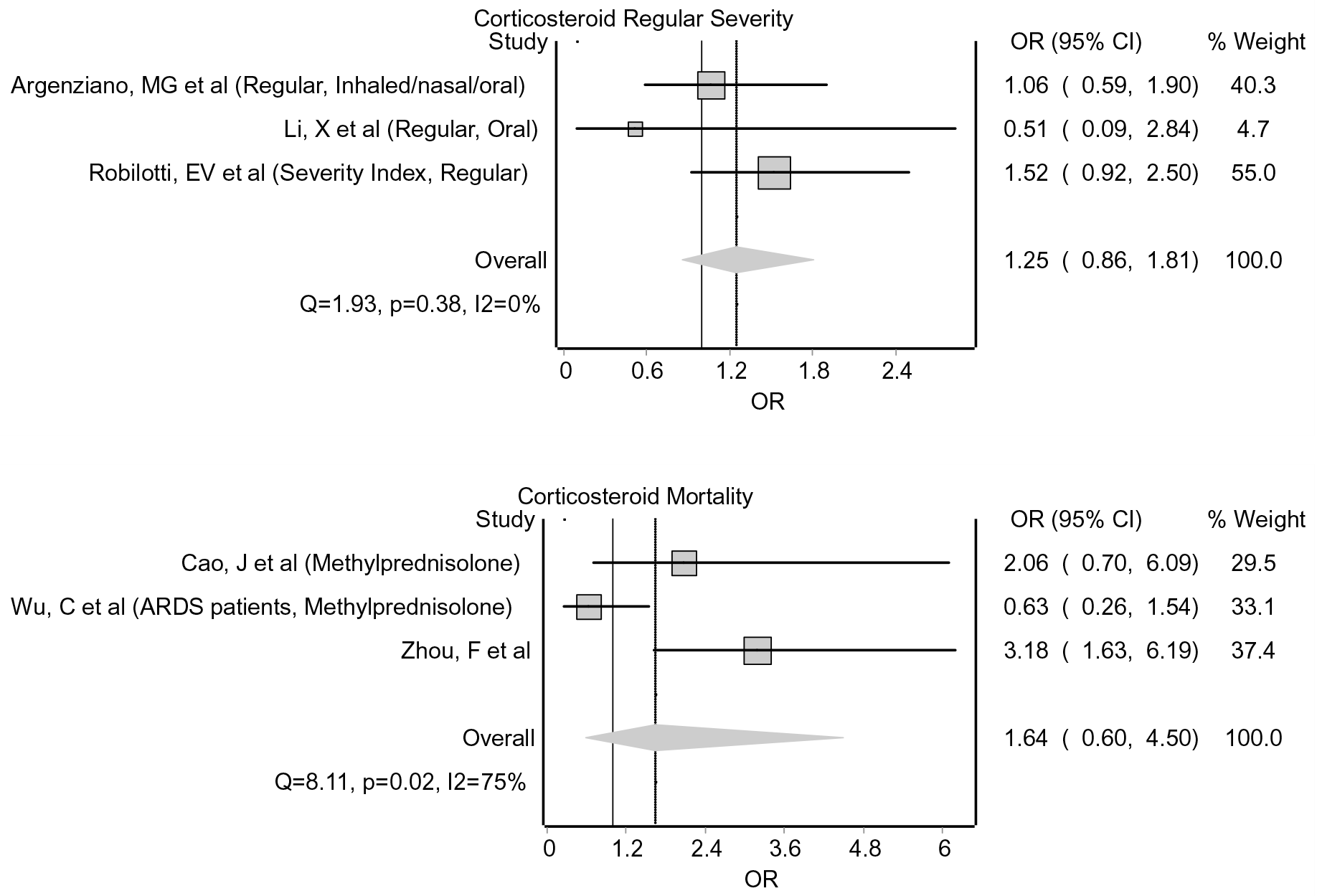

### Immunosuppressants

Susceptibility with immunosuppressants had one study not peer reviewed (Rentsch CT et al.) and sensitivity analysis was run for this already within the study.

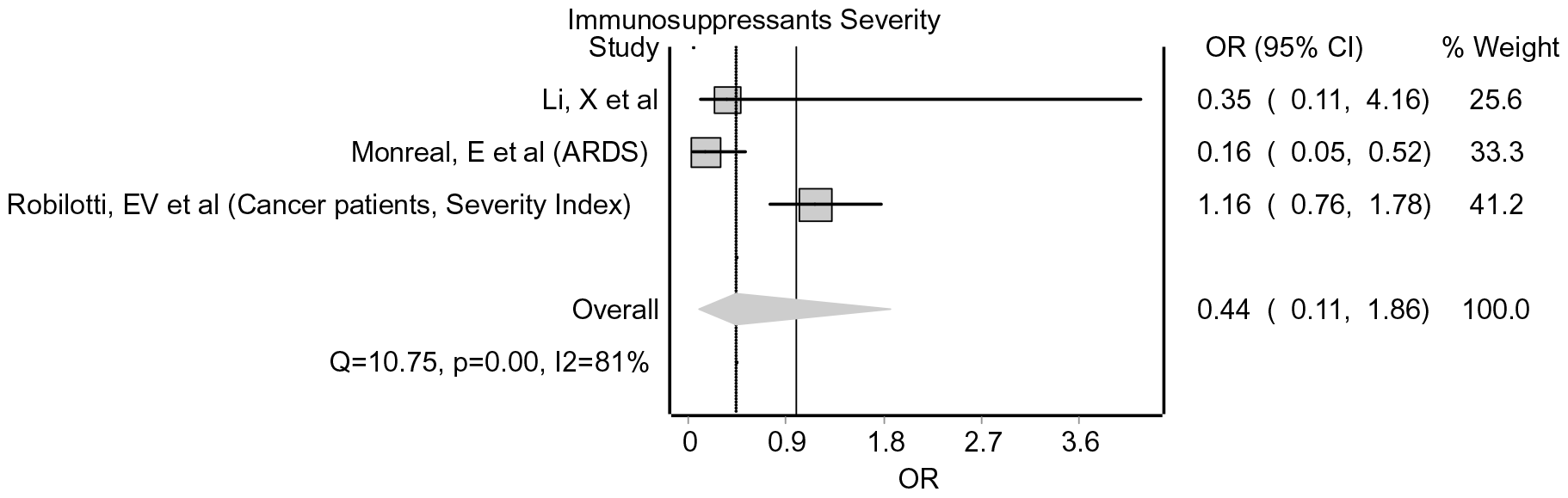

No meta-analysis was performed for mortality related to immunosuppressants.

### NSAIDs, statins, TZDs

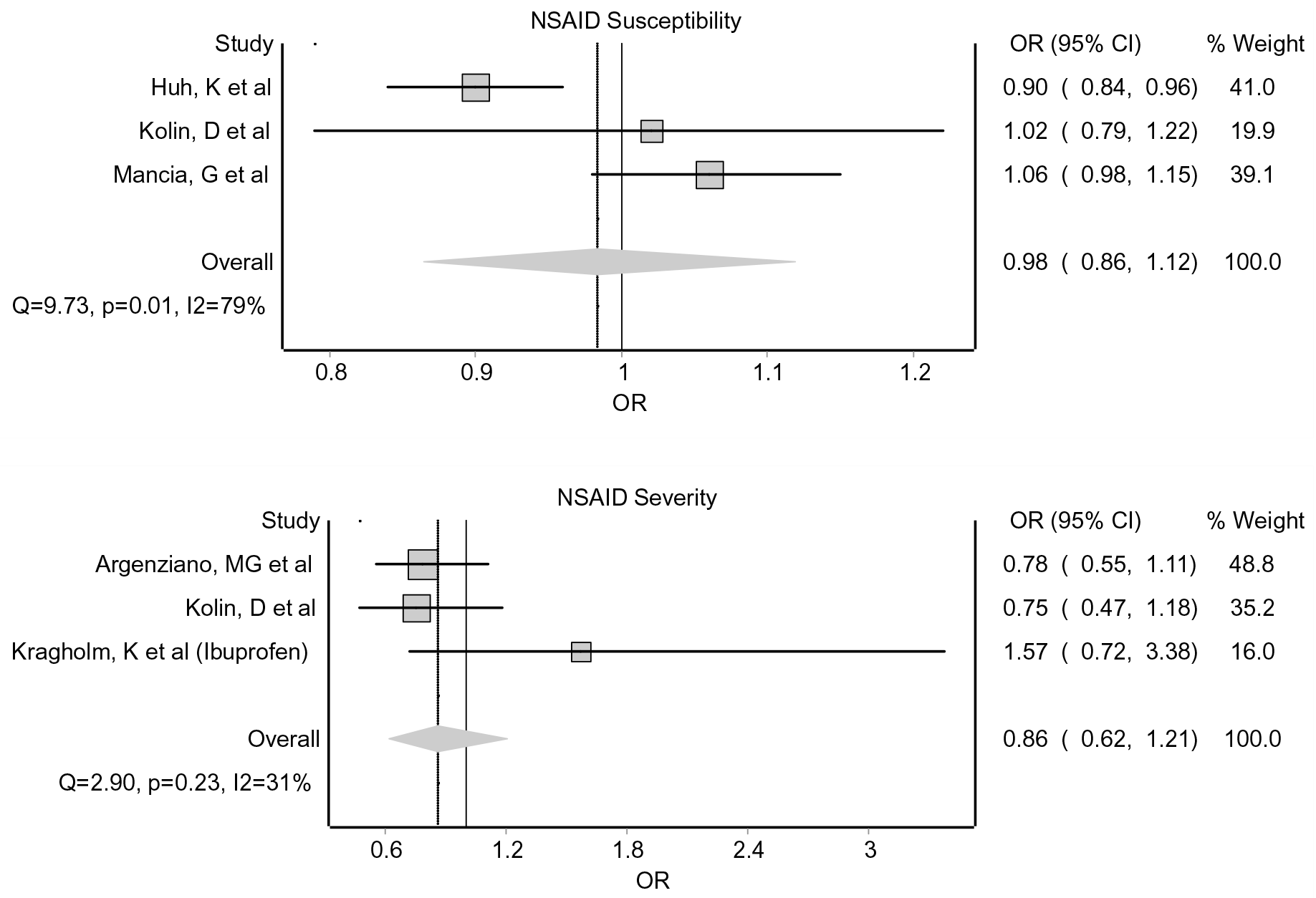

No meta-analyses were performed for mortality related to either NSAIDs, statins or TZDs.

No meta-analyses were performed for severity related to either statins or TZDs.

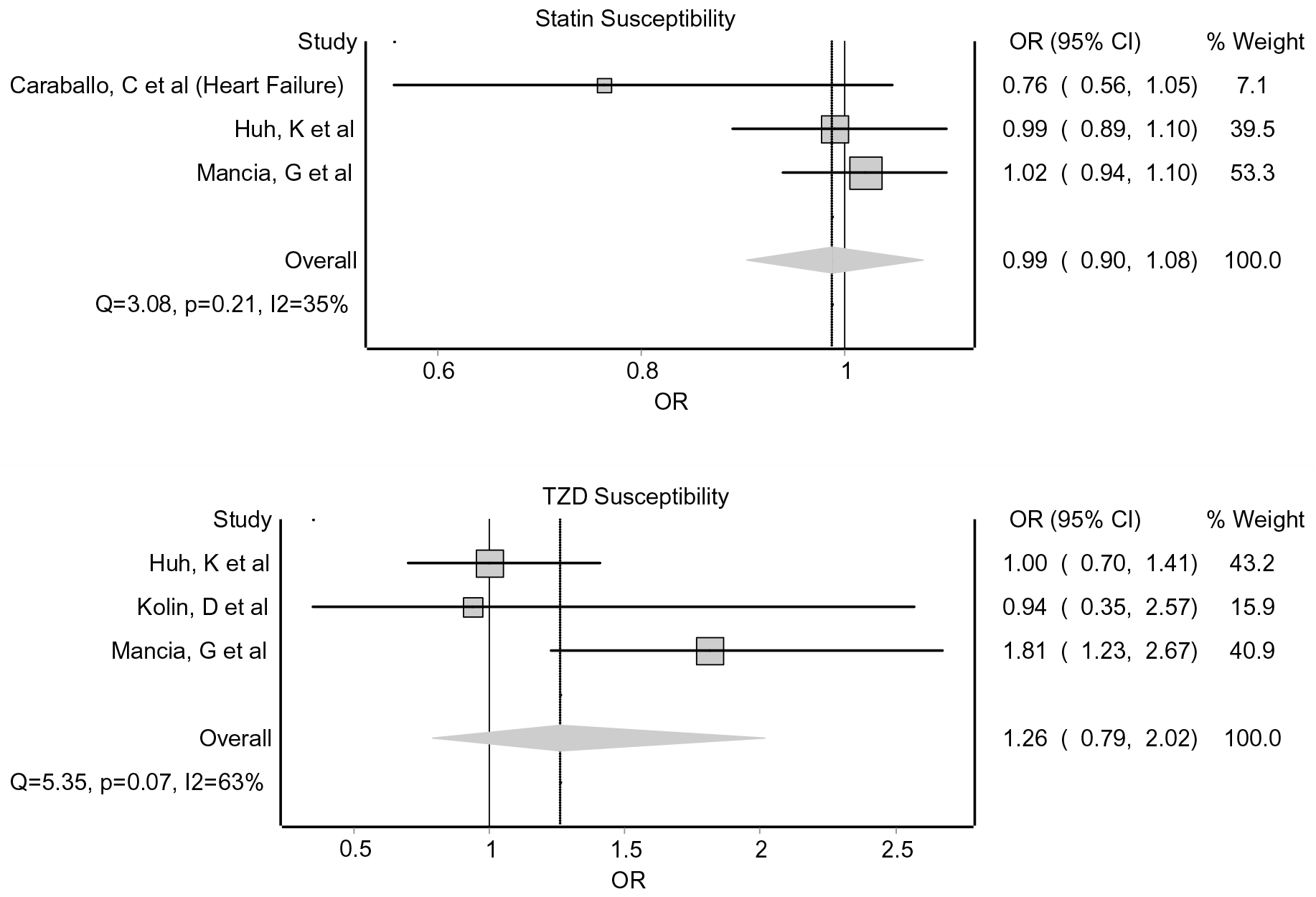

No meta-analyses were performed for MCRAs.

